# CLINICAL AND SEROLOGICAL PREDICTORS OF POST COVID-19 CONDITION – FINDINGS FROM A CANADIAN PROSPECTIVE COHORT STUDY

**DOI:** 10.1101/2023.07.29.23293334

**Authors:** Erin Collins, Yannick Galipeau, Corey Arnold, Anne Bhéreur, Ronald Booth, Arianne C. Buchan, Curtis Cooper, Angela M. Crawley, Pauline S. McCluskie, Michaeline McGuinty, Martin Pelchat, Lynda Rocheleau, Raphael Saginur, Chris Gravel, Steven Hawken, Marc-André Langlois, Julian Little

## Abstract

**Introduction:** More than three years into the pandemic, there is persisting uncertainty as to the etiology, biomarkers, and risk factors of Post COVID-19 Condition (PCC). Serological research data remain a largely untapped resource. Few studies have investigated the potential relationships between post-acute serology and PCC, while accounting for clinical covariates.

**Methods:** We compared clinical and serological predictors among COVID-19 survivors with (n=102 cases) and without (n=122 controls) persistent symptoms ≥12 weeks post-infection. We selected four primary serological predictors (anti-nucleocapsid (N), anti-Spike, and anti-receptor binding domain (RBD) IgG titres, and neutralization efficiency), and specified clinical covariates a priori.

**Results:** Similar proportions of PCC-cases (66.7%, n=68) and infected-controls (71.3%, n=87) tested positive for anti-N IgG. More cases tested positive for anti-Spike (94.1%, n=96) and anti-RBD (95.1%, n=97) IgG, as compared with controls (anti-Spike: 89.3%, n=109; anti-RBD: 84.4%, n=103). Similar trends were observed among unvaccinated participants. Effects of IgG titres on PCC status were non-significant in univariate and multivariate analyses. Adjusting for age and sex, PCC-cases were more likely to be efficient neutralizers (OR 2.2, 95% CI 1.11 – 4.49), and odds was further increased among cases to report deterioration in quality of life (OR 3.4, 95% CI 1.64 – 7.31). Clinical covariates found to be significantly related to PCC included obesity (OR 2.3, p=0.02), number of months post COVID-19 (OR 1.1, p<0.01), allergies (OR 1.8, p=0.04), and need for medical support (OR 4.1, p<0.01).

**Conclusion:** Despite past COVID-19 infection, approximately one third of PCC-cases and infected-controls were seronegative for anti-N IgG. Findings suggest higher neutralization efficiency among cases as compared with controls, and that this relationship is stronger among cases with more severe PCC. Cases also required more medical support for COVID-19 symptoms, and described complex, ongoing health sequelae. More data from larger cohorts are needed to substantiate results, permit subgroup analyses of IgG titres, and explore for differences between clusters of PCC symptoms. Future assessment of IgG subtypes may also elucidate new findings.

## Introduction

Post COVID-19 Condition (PCC), also known as Long COVID or Post-acute sequelae of COVID-19 (PASC), is a major public health concern with severe and pervasive impacts on physical and mental health [1–6]. More than three years into the pandemic, continued disparities as to the definition, presumed etiology, and prevalence of PCC deter efforts to detect and manage this condition [6–8]. Recent findings suggest that 10-20% of adults infected by COVID-19 will develop long-term symptoms [4,6,9]. These estimates tend to be higher in studies on patients hospitalized in the acute phase of illness and/or preceding the Omicron era [4,6,9,10]. Also, knowledge of how COVID-19 vaccination and infection/reinfection by SARS-CoV-2 Variants of Concern (VOCs) impact PCC onset, manifestations, and longevity continues to evolve [1,11–14]. There is an enduring need for rigorous, interdisciplinary efforts to examine multi-domain risk and protective factors of PCC onset, severity, and longevity, and its impacts on the healthcare system and economy [3–5,15].

Efforts to understand potential PCC predictors have largely relied on clinical records and self-reports of demographics, health history, and initial disease sequelae [16–18]. These studies have highlighted a number of potential clinical predictors (including age; sex; ethnicities; comorbidities, namely asthma, obesity, immune deficiency, lung disease, heart disease, kidney disease, and diabetes; severity, type, number, and duration of acute symptoms; need for hospitalization; lower socioeconomic status; stress; allergies; and smoking) [5,6,8,17–19], but are often bereft of variations in humoral response profile, which may be driven by severity and trajectory of COVID-19 infection and sequelae. There is a need to explore whether the inclusion of serological data can improve prediction of PCC, as compared with models based solely on clinical predictors. Also, if people with PCC are less likely to elicit a robust and sustained serological response post-infection, as compared to people without PCC, a blind reliance on serological evidence to diagnose COVID-19 infection may lead to underestimates of prevalence, and potentially exclude many people with PCC from participation in research studies, and qualifying for access to needed supports and services [3,5,16,20–21].

Studies on COVID-19 survivors comparing post-acute serological response between those with and without persistent symptoms have yielded highly mixed results [16,20,22–29]. Some reports suggest that people with PCC are more likely to have lower titres post-infection, as compared with survivors of COVID-19 without persistent symptoms. Non-detectable levels of antibodies post-infection may indicate need for testing strategies other than serological analysis. For example, Krishna *et al.* found evidence of persistent SARS-CoV-2-specific T cell responses in seronegative (negative for both anti-Spike and anti-N SARS-CoV-2 IgG) patients with PCC [21]. Conversely, other studies have found higher post-infection antibody titres to be associated with PCC, or no difference in humoral response in relation to persistent symptoms. Interpretation of these conflicting results is further complicated by differences in study populations, sample sizes, type of assay, number and type/subtype of target antigens, collection procedures, timing of sampling, and definition/assessment of PCC [5].

In this report, we summarize baseline findings for people found to have previous COVID-19 infection in a large Canadian prospective cohort study. We aimed to 1) describe clinical and serological characteristics among those with and without symptoms persisting ≥12 weeks post COVID-19, and 2) estimate associations between serological markers and PCC, accounting for clinical covariates. Among PCC-cases, we also described symptom characteristics, severity, and impact on quality of life.

## Materials & Methods

### Study population

The present analysis relates to a subgroup of participants from the Stop the Spread Ottawa (SSO) cohort study. Briefly, the SSO study on COVID-19 immune response recruited over 1000 adults in the Ottawa region from September 2020 to September 2021. All adults ≥18 years of age in the Ottawa region (1) at heightened risk of COVID-19 exposure/infection due to occupation or health condition, or (2) with any history of COVID-19 infection, confirmed by positive PCR test and/or serology, were eligible to participate. Starting in October 2020, participants provided monthly blood and saliva samples over a 10-month period. Enrolment closed September 2021. Conduct of this study was reviewed and approved by The Ottawa Health Science Network Hospital Research Ethics Board (2020-0481). All participants provided informed and written consent.

### Selection of PCC-cases and infected-controls

All SSO participants who reported a pre-baseline positive PCR test (external to study) and/or tested positive by serology at baseline (signal-to-cutoff ratio – S/CO ≥1.0 for anti-N IgG and S/CO ≥1.0 for either anti-S IgG or anti-RBD IgG) were considered for inclusion. Further inclusion criteria were that participants had contributed ≥ 1 blood specimen, and been assessed for persistent symptoms ≥12 weeks post initial positive PCR test (or, in the absence of positive test, due to which infection date could not be discerned, ≥12 weeks post baseline visit).

Participants meeting these inclusion criteria were defined to be PCC-cases if they reported any persistent symptoms, or infected-controls if they reported no persistent symptoms ≥12 weeks post-positive COVID-19 test/baseline visit. Participants who reported the presence or absence of persistent symptoms <12 weeks post-infection and then left the study were excluded. Though we included participants regardless of vaccination status, less than a quarter of participants (23.2%, n=52) received ≥1 COVID-19 vaccines ≥14 days prior to baseline visit, and few participants (2.2%, n=5) received ≥1 vaccines ≥14 days prior to COVID-19 infection date.

### Serological predictors

At baseline, one (5 mL) tube with a separator gel with clot activator for serum and two (10 mL × 2) tubes with EDTA for lymphocyte isolation were drawn. Serological testing included main isotypes IgA, IgM, IgG against COVID-19 N, RBD, and Spike antigens. Neutralizing efficiency against the SARS-CoV-2 Spike protein was also assessed. Full methods were published previously [30].

We examined the relationship between PCC and 1) anti-Spike, anti-N, and anti-RBD IgG titres (scaled luminescent units - SLU); and 2) % efficient neutralizers (≥85% inhibition against SARS-CoV-2 Spike protein). The cut-off for neutralization efficiency was determined by the study team to develop the in-house made surrogate neutralization enzyme-linked immunosorbent assay (snELISA) used for SSO serological analysis [31].

### Collection of data on clinical covariates and PCC descriptors

Participants in the study responded to questions at baseline, and three- and 10-months post baseline via an electronic survey with the following categories: demographics and health history; severity of COVID-19 signs and symptoms; risks of exposure; and socioeconomic and psychosocial impacts of the pandemic. Responses in all categories were compared between PCC-cases and infected-controls. All participants were asked to complete the 10-item Kessler Psychological Distress Scale (K10), an internationally validated tool for the screening and assessment of psychological distress [32]. Participants who reported a pre-baseline positive PCR test also completed the 15-item Impact of Event Scale (IES), used to assess for post-traumatic stress (PTS) symptoms [33].

Dates of COVID-19 positive tests and vaccines were self-reported by participants and verified where possible through use of medical and laboratory records, including Eastern Ontario Regional Laboratory Association (EORLA) reports and The Ottawa Hospital COVID-19 Registry to identify SARS-CoV-2. We reported the small numbers of PCC-cases and infected-controls without any lab-confirmed evidence of past infection (where self-reported PCR test could not be verified by laboratory record, and with negative serology at baseline).

Participants reporting persistent symptoms at time of survey completion were asked to self-rate severity of chronic symptoms, and impact of symptoms on quality of life (QoL). PCC descriptors (symptom type and severity of symptoms) and need for healthcare/medical supports were compared between cases who did and did not report worsening QoL due to PCC symptoms.

### Statistical analysis

Descriptive analyses included frequency tables (categorical) and measures of centre or spread (continuous). Bivariate analyses were conducted between each predictor and outcome, and among predictors using the chi-square/Fisher’s exact test (categorical) and Wilcoxon-Rank Sum test (continuous) at alpha level 0.05. We constructed a series of logistic regression models to assess the relationship between each primary serological predictor (anti-Spike, anti-N, and anti-RBD IgG levels, and neutralizing efficiency) and PCC, respectively. All covariates considered for inclusion were identified a priori. We assessed each model for confounding, collinearity, and outliers. Given few missing data (<5% per variable), we decided to use complete case analysis. In each model, we tested for interactions between the primary predictor, and sex or time post-infection. Age, number of months post COVID-19, and IgG titres were fit with restricted cubic splines (RCS, 3 knots at the 10th, 50th and 90th percentiles) to account for potential non-linearities [34], using the rms R package. For each primary predictor, we compared a minimally adjusted model (age and sex) with the best performing fully adjusted model. We used the Bayesian information criterion (BIC) to inform the selection of covariates for the fully adjusted model. We reported unadjusted and adjusted odds ratios with 95% confidence intervals (CIs) and goodness-of-fit using the C-statistic [34]. We presented receiver operating characteristic (ROC) curves and plotted results for RCS-transformed predictors by sex. We used sensitivity analyses to assess effects from removing 1) outliers, 2) receipt of ≥ 1 pre-infection COVID-19 vaccines; 3) receipt of ≥ 2 pre-baseline COVID-19 vaccines, and 4) baseline serology collected <14 days or >365 days post COVID-19 infection, or where days post COVID-19 could not be determined given no record of infection prior to study blood sampling. All analyses were conducted with SAS 9.4 and R, 4.2.1.

## Results

### Comparisons of serological and clinical predictors

#### Demographics and health history

We identified 102 PCC-cases and 122 infected-controls meeting study criteria. **Figure 1** displays the selection procedures of study participants. Baseline characteristics are summarized in **Table 1**. Participants ranged in age from 21-75 years old. There were less males among PCC-cases (35.3%, n=36), than infected-controls (42.6%, n=52). Few participants were non-white (12.1%, n=27). Over 75% of participants were employed at time of baseline visit (n=176), and about 26% (n=58) reported annual household income before taxes as $150,000 or more. PCC-cases had higher rates of pre COVID-19 obesity (25.5%, n=26); asthma (13.7%, n=14); and diabetes (8.8%, n=9), as compared to infected-controls (obesity – 13.1%, n=16; asthma – 6.6%, n=8; diabetes – 4.1%, n=5).

**FIGURE 1:**
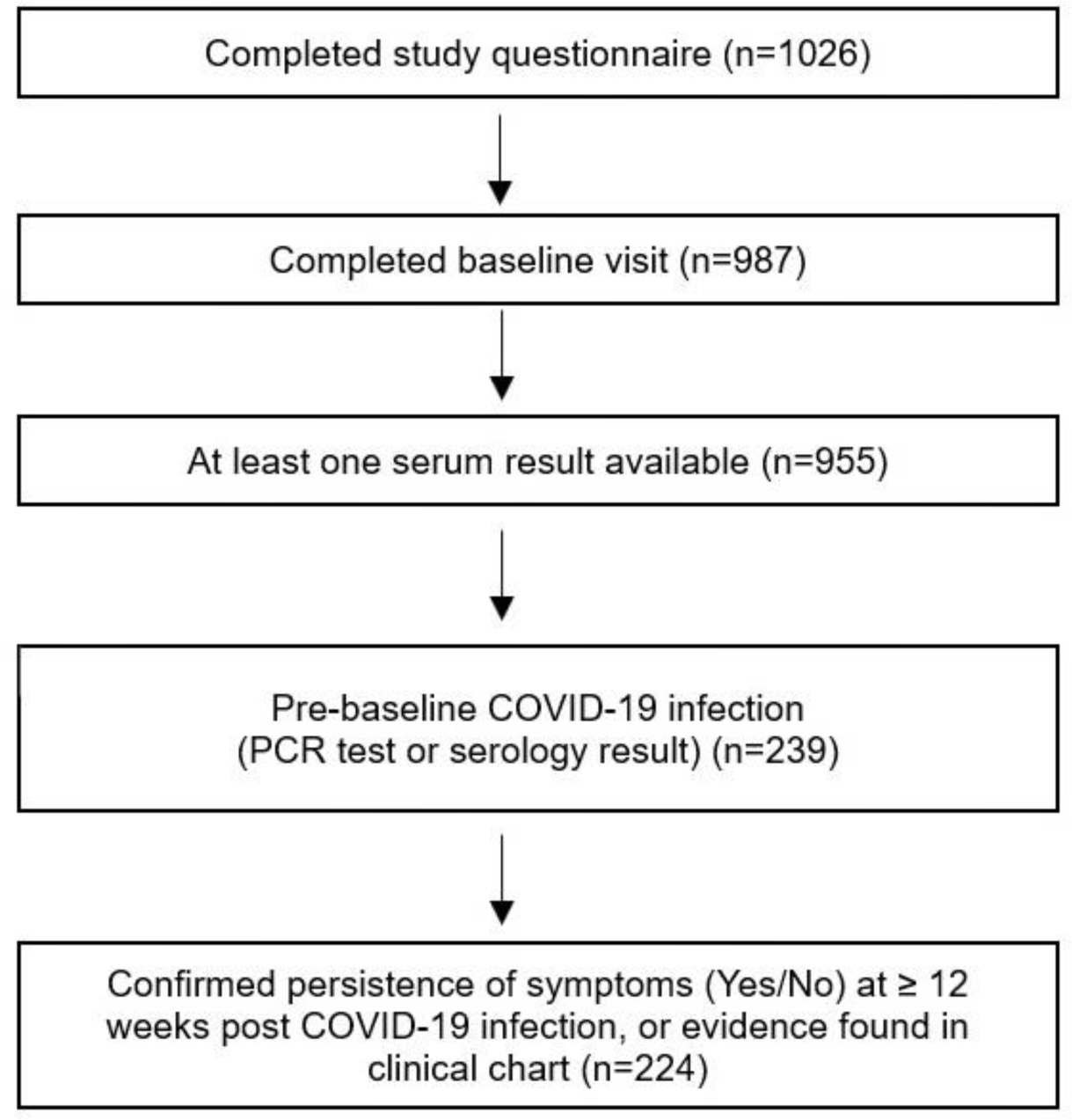
Selection of study participants from the Stop the Spread Ottawa study.

**TABLE 1:**
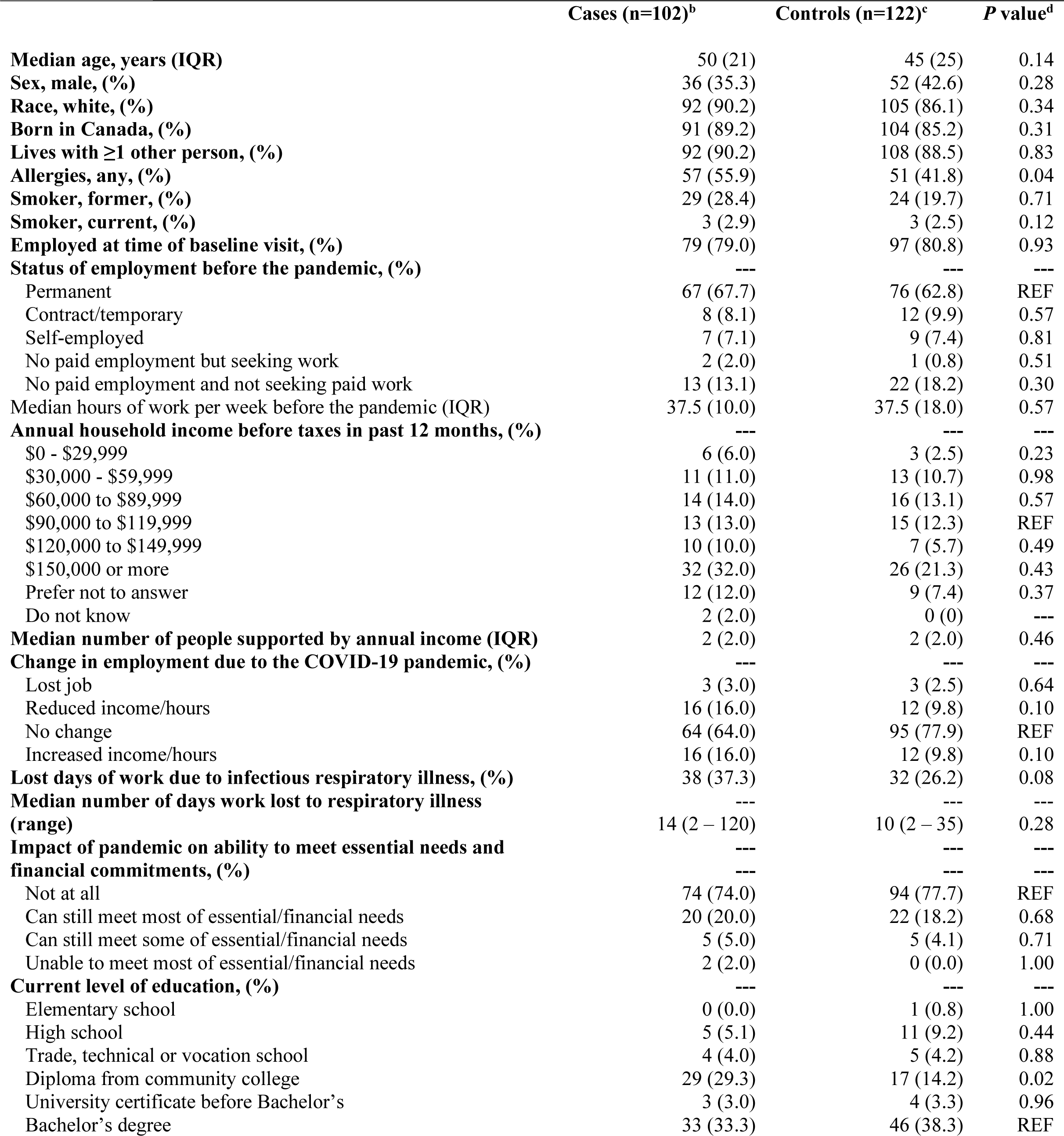
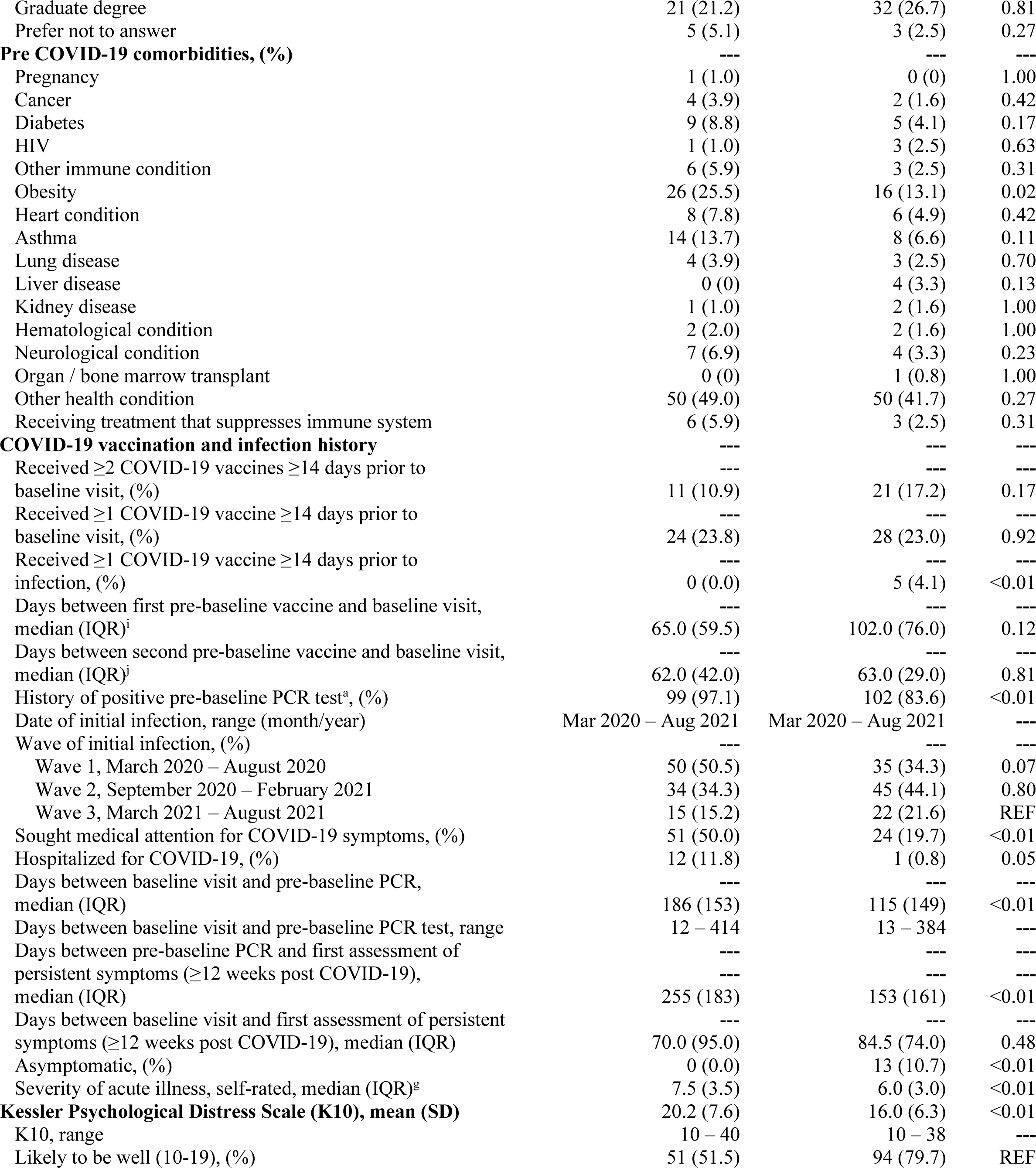
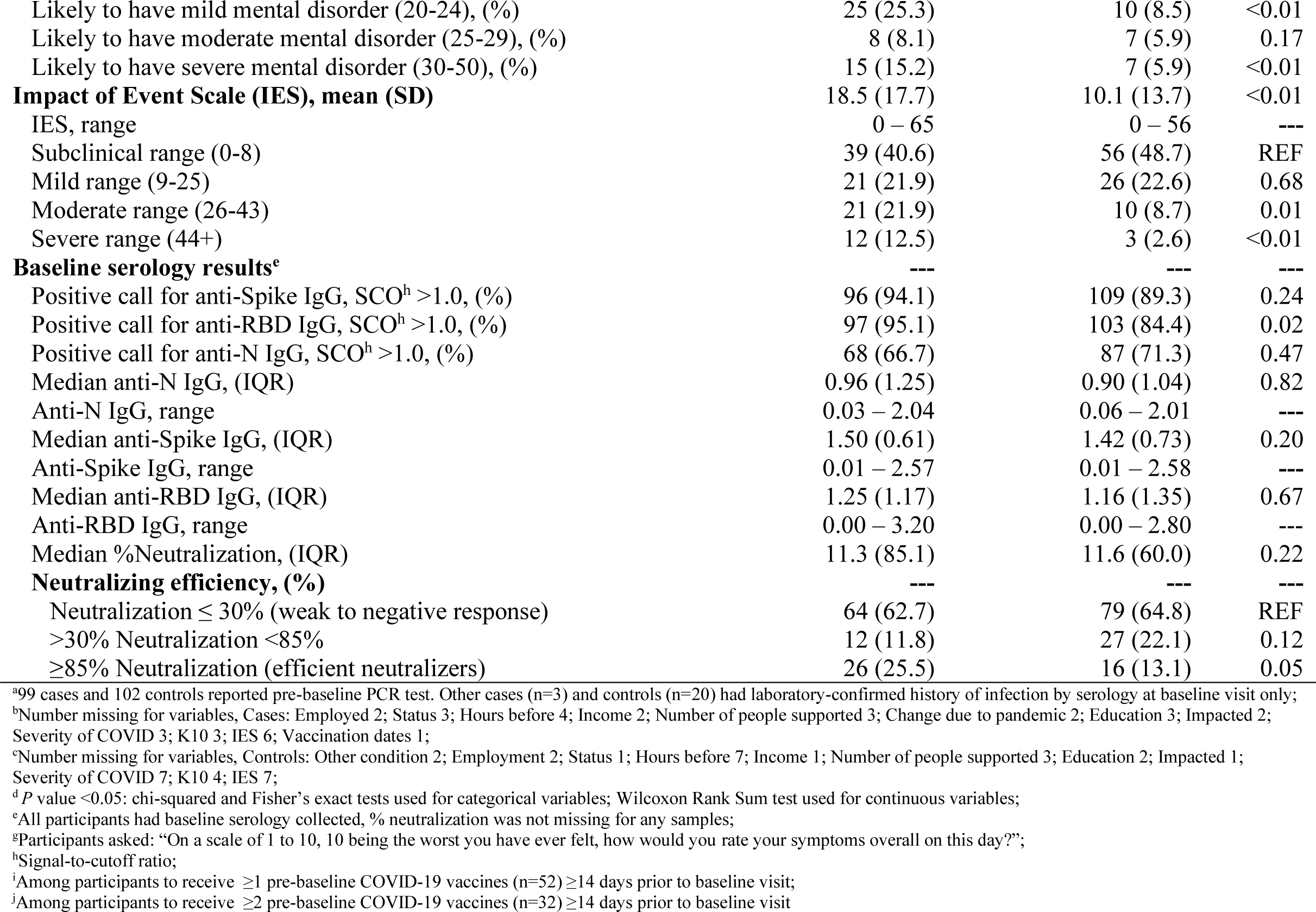
Baseline characteristics of PCC-cases with symptoms persisting ≥12 weeks and infected-controls without persistent symptoms.

|  | Cases (n=102)<sup>b</sup> | Controls (n=122)<sup>c</sup> | P value<sup>d</sup> |
| --- | --- | --- | --- |
| **Median age, years (IQR)** | 50 (21) | 45 (25) | 0.14 |
| **Sex, male, (%)** | 36 (35.3) | 52 (42.6) | 0.28 |
| **Race, white, (%)** | 92 (90.2) | 105 (86.1) | 0.34 |
| **Born in Canada, (%)** | 91 (89.2) | 104 (85.2) | 0.31 |
| **Lives with $\geq 1$ other person, (%)** | 92 (90.2) | 108 (88.5) | 0.83 |
| **Allergies, any, (%)** | 57 (55.9) | 51 (41.8) | 0.04 |
| **Smoker, former, (%)** | 29 (28.4) | 24 (19.7) | 0.71 |
| **Smoker, current, (%)** | 3 (2.9) | 3 (2.5) | 0.12 |
| **Employed at time of baseline visit, (%)** | 79 (79.0) | 97 (80.8) | 0.93 |
| **Status of employment before the pandemic, (%)** | --- | --- | --- |
| Permanent | 67 (67.7) | 76 (62.8) | REF |
| Contract/temporary | 8 (8.1) | 12 (9.9) | 0.57 |
| Self-employed | 7 (7.1) | 9 (7.4) | 0.81 |
| No paid employment but seeking work | 2 (2.0) | 1 (0.8) | 0.51 |
| No paid employment and not seeking paid work | 13 (13.1) | 22 (18.2) | 0.30 |
| **Median hours of work per week before the pandemic (IQR)** | 37.5 (10.0) | 37.5 (18.0) | 0.57 |
| **Annual household income before taxes in past 12 months, (%)** | --- | --- | --- |
| \$0 - \$29,999 | 6 (6.0) | 3 (2.5) | 0.23 |
| \$30,000 - \$59,999 | 11 (11.0) | 13 (10.7) | 0.98 |
| \$60,000 to \$89,999 | 14 (14.0) | 16 (13.1) | 0.57 |
| \$90,000 to \$119,999 | 13 (13.0) | 15 (12.3) | REF |
| \$120,000 to \$149,999 | 10 (10.0) | 7 (5.7) | 0.49 |
| \$150,000 or more | 32 (32.0) | 26 (21.3) | 0.43 |
| Prefer not to answer | 12 (12.0) | 9 (7.4) | 0.37 |
| Do not know | 2 (2.0) | 0 (0) | --- |
| **Median number of people supported by annual income (IQR)** | 2 (2.0) | 2 (2.0) | 0.46 |
| **Change in employment due to the COVID-19 pandemic, (%)** | --- | --- | --- |
| Lost job | 3 (3.0) | 3 (2.5) | 0.64 |
| Reduced income/hours | 16 (16.0) | 12 (9.8) | 0.10 |
| No change | 64 (64.0) | 95 (77.9) | REF |
| Increased income/hours | 16 (16.0) | 12 (9.8) | 0.10 |
| **Lost days of work due to infectious respiratory illness, (%)** | 38 (37.3) | 32 (26.2) | 0.08 |
| **Median number of days work lost to respiratory illness (range)** | 14 (2 – 120) | 10 (2 – 35) | 0.28 |
| **Impact of pandemic on ability to meet essential needs and financial commitments, (%)** | --- | --- | --- |
| Not at all | 74 (74.0) | 94 (77.7) | REF |
| Can still meet most of essential/financial needs | 20 (20.0) | 22 (18.2) | 0.68 |
| Can still meet some of essential/financial needs | 5 (5.0) | 5 (4.1) | 0.71 |
| Unable to meet most of essential/financial needs | 2 (2.0) | 0 (0.0) | 1.00 |
| **Current level of education, (%)** | --- | --- | --- |
| Elementary school | 0 (0.0) | 1 (0.8) | 1.00 |
| High school | 5 (5.1) | 11 (9.2) | 0.44 |
| Trade, technical or vocation school | 4 (4.0) | 5 (4.2) | 0.88 |
| Diploma from community college | 29 (29.3) | 17 (14.2) | 0.02 |
| University certificate before Bachelor's | 3 (3.0) | 4 (3.3) | 0.96 |
| Bachelor's degree | 33 (33.3) | 46 (38.3) | REF |
| Graduate degree | 21 (21.2) | 32 (26.7) | 0.81 |
| Prefer not to answer | 5 (5.1) | 3 (2.5) | 0.27 |
| **Pre COVID-19 comorbidities, (%)** | --- | --- | --- |
| Pregnancy | 1 (1.0) | 0 (0) | 1.00 |
| Cancer | 4 (3.9) | 2 (1.6) | 0.42 |
| Diabetes | 9 (8.8) | 5 (4.1) | 0.17 |
| HIV | 1 (1.0) | 3 (2.5) | 0.63 |
| Other immune condition | 6 (5.9) | 3 (2.5) | 0.31 |
| Obesity | 26 (25.5) | 16 (13.1) | 0.02 |
| Heart condition | 8 (7.8) | 6 (4.9) | 0.42 |
| Asthma | 14 (13.7) | 8 (6.6) | 0.11 |
| Lung disease | 4 (3.9) | 3 (2.5) | 0.70 |
| Liver disease | 0 (0) | 4 (3.3) | 0.13 |
| Kidney disease | 1 (1.0) | 2 (1.6) | 1.00 |
| Hematological condition | 2 (2.0) | 2 (1.6) | 1.00 |
| Neurological condition | 7 (6.9) | 4 (3.3) | 0.23 |
| Organ / bone marrow transplant | 0 (0) | 1 (0.8) | 1.00 |
| Other health condition | 50 (49.0) | 50 (41.7) | 0.27 |
| Receiving treatment that suppresses immune system | 6 (5.9) | 3 (2.5) | 0.31 |
| **COVID-19 vaccination and infection history** | --- | --- | --- |
| Received $\geq 2$ COVID-19 vaccines $\geq 14$ days prior to baseline visit, (%) | 11 (10.9) | 21 (17.2) | 0.17 |
| Received $\geq 1$ COVID-19 vaccine $\geq 14$ days prior to baseline visit, (%) | 24 (23.8) | 28 (23.0) | 0.92 |
| Received $\geq 1$ COVID-19 vaccine $\geq 14$ days prior to infection, (%) | 0 (0.0) | 5 (4.1) | <0.01 |
| Days between first pre-baseline vaccine and baseline visit, median (IQR)<sup>i</sup> | 65.0 (59.5) | 102.0 (76.0) | 0.12 |
| Days between second pre-baseline vaccine and baseline visit, median (IQR)<sup>j</sup> | 62.0 (42.0) | 63.0 (29.0) | 0.81 |
| History of positive pre-baseline PCR test<sup>a</sup>, (%) | 99 (97.1) | 102 (83.6) | <0.01 |
| Date of initial infection, range (month/year) | Mar 2020 – Aug 2021 | Mar 2020 – Aug 2021 | --- |
| Wave of initial infection, (%) | --- | --- | --- |
| Wave 1, March 2020 – August 2020 | 50 (50.5) | 35 (34.3) | 0.07 |
| Wave 2, September 2020 – February 2021 | 34 (34.3) | 45 (44.1) | 0.80 |
| Wave 3, March 2021 – August 2021 | 15 (15.2) | 22 (21.6) | REF |
| Sought medical attention for COVID-19 symptoms, (%) | 51 (50.0) | 24 (19.7) | <0.01 |
| Hospitalized for COVID-19, (%) | 12 (11.8) | 1 (0.8) | 0.05 |
| Days between baseline visit and pre-baseline PCR, median (IQR) | 186 (153) | 115 (149) | <0.01 |
| Days between baseline visit and pre-baseline PCR test, range | 12 – 414 | 13 – 384 | --- |
| Days between pre-baseline PCR and first assessment of persistent symptoms ($\geq 12$ weeks post COVID-19), median (IQR) | 255 (183) | 153 (161) | <0.01 |
| Days between baseline visit and first assessment of persistent symptoms ($\geq 12$ weeks post COVID-19), median (IQR) | 70.0 (95.0) | 84.5 (74.0) | 0.48 |
| Asymptomatic, (%) | 0 (0.0) | 13 (10.7) | <0.01 |
| Severity of acute illness, self-rated, median (IQR)<sup>g</sup> | 7.5 (3.5) | 6.0 (3.0) | <0.01 |
| **Kessler Psychological Distress Scale (K10), mean (SD)** | 20.2 (7.6) | 16.0 (6.3) | <0.01 |
| K10, range | 10 – 40 | 10 – 38 | --- |
| Likely to be well (10-19), (%) | 51 (51.5) | 94 (79.7) | REF |
| Likely to have mild mental disorder (20-24), (%) | 25 (25.3) | 10 (8.5) | <0.01 |
| Likely to have moderate mental disorder (25-29), (%) | 8 (8.1) | 7 (5.9) | 0.17 |
| Likely to have severe mental disorder (30-50), (%) | 15 (15.2) | 7 (5.9) | <0.01 |
| **Impact of Event Scale (IES), mean (SD)** | 18.5 (17.7) | 10.1 (13.7) | <0.01 |
| IES, range | 0 – 65 | 0 – 56 | --- |
| Subclinical range (0-8) | 39 (40.6) | 56 (48.7) | REF |
| Mild range (9-25) | 21 (21.9) | 26 (22.6) | 0.68 |
| Moderate range (26-43) | 21 (21.9) | 10 (8.7) | 0.01 |
| Severe range (44+) | 12 (12.5) | 3 (2.6) | <0.01 |
| **Baseline serology results<sup>e</sup>** | --- | --- | --- |
| Positive call for anti-Spike IgG, SCO<sup>h</sup> >1.0, (%) | 96 (94.1) | 109 (89.3) | 0.24 |
| Positive call for anti-RBD IgG, SCO<sup>h</sup> >1.0, (%) | 97 (95.1) | 103 (84.4) | 0.02 |
| Positive call for anti-N IgG, SCO<sup>h</sup> >1.0, (%) | 68 (66.7) | 87 (71.3) | 0.47 |
| Median anti-N IgG, (IQR) | 0.96 (1.25) | 0.90 (1.04) | 0.82 |
| Anti-N IgG, range | 0.03 – 2.04 | 0.06 – 2.01 | --- |
| Median anti-Spike IgG, (IQR) | 1.50 (0.61) | 1.42 (0.73) | 0.20 |
| Anti-Spike IgG, range | 0.01 – 2.57 | 0.01 – 2.58 | --- |
| Median anti-RBD IgG, (IQR) | 1.25 (1.17) | 1.16 (1.35) | 0.67 |
| Anti-RBD IgG, range | 0.00 – 3.20 | 0.00 – 2.80 | --- |
| Median %Neutralization, (IQR) | 11.3 (85.1) | 11.6 (60.0) | 0.22 |
| **Neutralizing efficiency, (%)** | --- | --- | --- |
| Neutralization ≤ 30% (weak to negative response) | 64 (62.7) | 79 (64.8) | REF |
| >30% Neutralization <85% | 12 (11.8) | 27 (22.1) | 0.12 |
| ≥85% Neutralization (efficient neutralizers) | 26 (25.5) | 16 (13.1) | 0.05 |

<sup>a</sup>99 cases and 102 controls reported pre-baseline PCR test. Other cases (n=3) and controls (n=20) had laboratory-confirmed history of infection by serology at baseline visit only;
<sup>b</sup>Number missing for variables, Cases: Employed 2; Status 3; Hours before 4; Income 2; Number of people supported 3; Change due to pandemic 2; Education 3; Impacted 2; Severity of COVID 3; K10 3; IES 6; Vaccination dates 1;
<sup>c</sup>Number missing for variables, Controls: Other condition 2; Employment 2; Status 1; Hours before 7; Income 1; Number of people supported 3; Education 2; Impacted 1; Severity of COVID 7; K10 4; IES 7;
<sup>d</sup>P value <0.05: chi-squared and Fisher's exact tests used for categorical variables; Wilcoxon Rank Sum test used for continuous variables;
<sup>e</sup>All participants had baseline serology collected, % neutralization was not missing for any samples;
<sup>f</sup>Participants asked: "On a scale of 1 to 10, 10 being the worst you have ever felt, how would you rate your symptoms overall on this day?";
<sup>h</sup>Signal-to-cutoff ratio;
<sup>i</sup>Among participants to receive ≥1 pre-baseline COVID-19 vaccines (n=52) ≥14 days prior to baseline visit;
<sup>j</sup>Among participants to receive ≥2 pre-baseline COVID-19 vaccines (n=32) ≥14 days prior to baseline visit

#### COVID-19 infection and vaccination history

Most participants reported a pre-baseline PCR test (89.7%, n=201), of which we were able to verify 92.5% (n=186) by laboratory or clinical record. Of the 7.5% (n=15) we were not able to verify, only two participants had negative serology at baseline. Participants who did not report a pre-baseline PCR test and had serological markers of infection at baseline (10.3%, n=23) were also included so long as they were assessed for symptoms persisting ≥12 weeks. Approximately 23% of PCC-cases (n=24) and infected-controls (n=28) received ≥1 vaccines ≥14 days prior to baseline visit, while 10.9% of cases (n=11) and 17.2% of controls (n=21) received ≥2 vaccines ≥14 days before baseline visit (**Table 1**). Results for unvaccinated and vaccinated participants are presented in **Tables S1a and S1b** respectively. Initial test dates ranged from March 2020 to August 2021. Few PCC-cases (11.8%, n=12) and infected-controls (0.8%, n=1) were hospitalized for COVID-19. However, half of PCC-cases (n=51) reported seeking medical attention for COVID-19 symptoms other than hospitalization, as compared to infected-controls (19.7%, n=24). Among controls, 10.7% (n=13) were asymptomatic while all cases had symptoms during acute illness. PCC-cases also self-rated the severity of overall symptoms higher (median of 7.5/10, IQR 3.5) on the worst day of acute illness, as compared to controls (median of 6.0/10 IQR 3.0). Participants to test positive by serology only, with no self-report of positive PCR test at baseline, were not asked to self-rate symptom severity and date of onset is often indeterminable. PCC-cases had a significantly longer follow-up time (median 186 days, IQR 183) between pre-baseline PCR and baseline serological sampling, as compared with infected-controls (median 115 days, IQR 149, p<0.01).

#### Anti-N, anti-Spike, and anti-RBD IgG titres

Similar proportions of cases (66.7%, n=68) and controls (71.3%, n=87, p=0.47) tested positive for anti-N IgG. Cases tended to have higher IgG titres (anti-N, anti-Spike, and anti-RBD), but differences were non-significant. Findings were similar upon restricting to unvaccinated participants (**Table S1a**) and varying post-infection time intervals (**Tables S2 a-c**, 14-365 days; 14-180 days; and 14-90 days post COVID-19 infection). Unvaccinated PCC-cases had higher anti-Spike IgG levels (median 1.45, IQR 0.79) than unvaccinated infected-controls (median 1.29, IQR 0.86, p=0.14). Among participants who attended baseline14-365 days, 14-180, or 14-90 days post-infection, IgG titres remained consistently higher among cases, except for anti-RBD assessed 14-90 days post-infection (PCC-cases – median anti-RBD 0.92, IQR 1.17; infected-controls – 1.03, IQR 1.40, p=0.04).

#### Neutralizing efficiency

More cases (25.5%, n=26) than controls (13.1%, n=16) were efficient neutralizers (≥85% neutralizing efficiency). As expected, most efficient neutralizers had received ≥1 COVID-19 vaccines prior to baseline serology sampling (**Table S1b**). However, median neutralization efficiency was somewhat higher among non-vaccinated cases (7.17, IQR 20.35), as compared to non-vaccinated controls (3.62, IQR 17.40, p=0.45), despite the former having a significantly longer median time interval (cases – 205, IQR 163 days; controls – 84, IQR 152, p<0.01) between COVID-19 onset and baseline visit (**Table S1a)**. Among 52 (23.2%) participants to receive ≥1 vaccines ≥14 days prior to baseline visit (**Table S1b**), cases had somewhat higher neutralizing efficiency (median 98.69, IQR 4.62), than controls (median 87.96, IQR 35.28, p=0.06). Similar trends were observed when limiting to different post-infection time intervals (**Tables S2 a-c**).

#### Socioeconomic and psychosocial impacts of the pandemic and COVID-19 infection

The majority of participants reported no change in employment or ability to meet essential needs due to the COVID-19 pandemic (**Table 1**). Differences in annual household income were non-significant. Somewhat more PCC-cases (37.3%, n=38) reported one or more lost days of work due to respiratory illness and a higher number of days lost (median 14, IQR 118), than infected-controls (median 10, IQR 33, p=0.28). Cases had a higher mean K10 (Kessler Psychological Distress Scale) score (20.2, SD 7.6) as compared to controls (16.0, SD 6.3, p<0.01), and more cases were likely to have mild (25.3%, n=25), moderate (8.1%, n=8), and severe (15.2%, n=15) mental disorder, than controls (mild disorder – 8.5%, n=10; moderate disorder – 5.9%, n=7; and severe disorder – 5.9%, n=7), using K10 cut-offs established previously [32]. PCC-cases also had a higher mean IES (Impact of Event Scale) score (18.5, SD 17.7) than infected-controls without PCC (10.1, SD 13.7, p<0.01). Applying previously applied IES cut-offs [33], more PCC-cases had moderate (21.9%, n=21) and severe (12.5%, n=12) distress than infected-controls (moderate distress – 8.7%, n=10; severe distress – 2.6%, n=3).

### Descriptors of Post COVID-19 Condition

**Table 2** delineates the number and type of persistent symptoms among all PCC-cases (n=102), those cases who reported worsened QoL post COVID-19 (n=65), and those who did not (n=37). The three most frequent symptoms described by PCC-cases with worsened QoL were fatigue (73.8%, n=48); shortness of breath (64.6%, n=42); and difficulties with thinking/concentrating (60.0%, n=39). Among PCC-cases who did not report worsened QoL (36.3%, n=37), the most common symptoms were loss of smell (n=14, 37.8%); loss of taste (n=9, 24.3%); and fatigue (n=12; 32.4%). The majority of cases (89.2%, n=91) reported ongoing symptoms for a median of 255 days (IQR 183; range 75-451 days) at time of first assessment for PCC. Among these cases, those who reported worsened QoL self-rated severity of persisting symptoms higher (median 4.0, IQR 4.0) than the other cases (median 2.0, IQR 2.0) on a scale of 1-10 (**Table 2**). PCC-cases endorsing worsened QoL also reported a higher number of post COVID-19 symptoms (median 6.0, IQR 9.0), than cases not reporting worsened QoL (median 2.0, IQR 3.0).

**TABLE 2:**
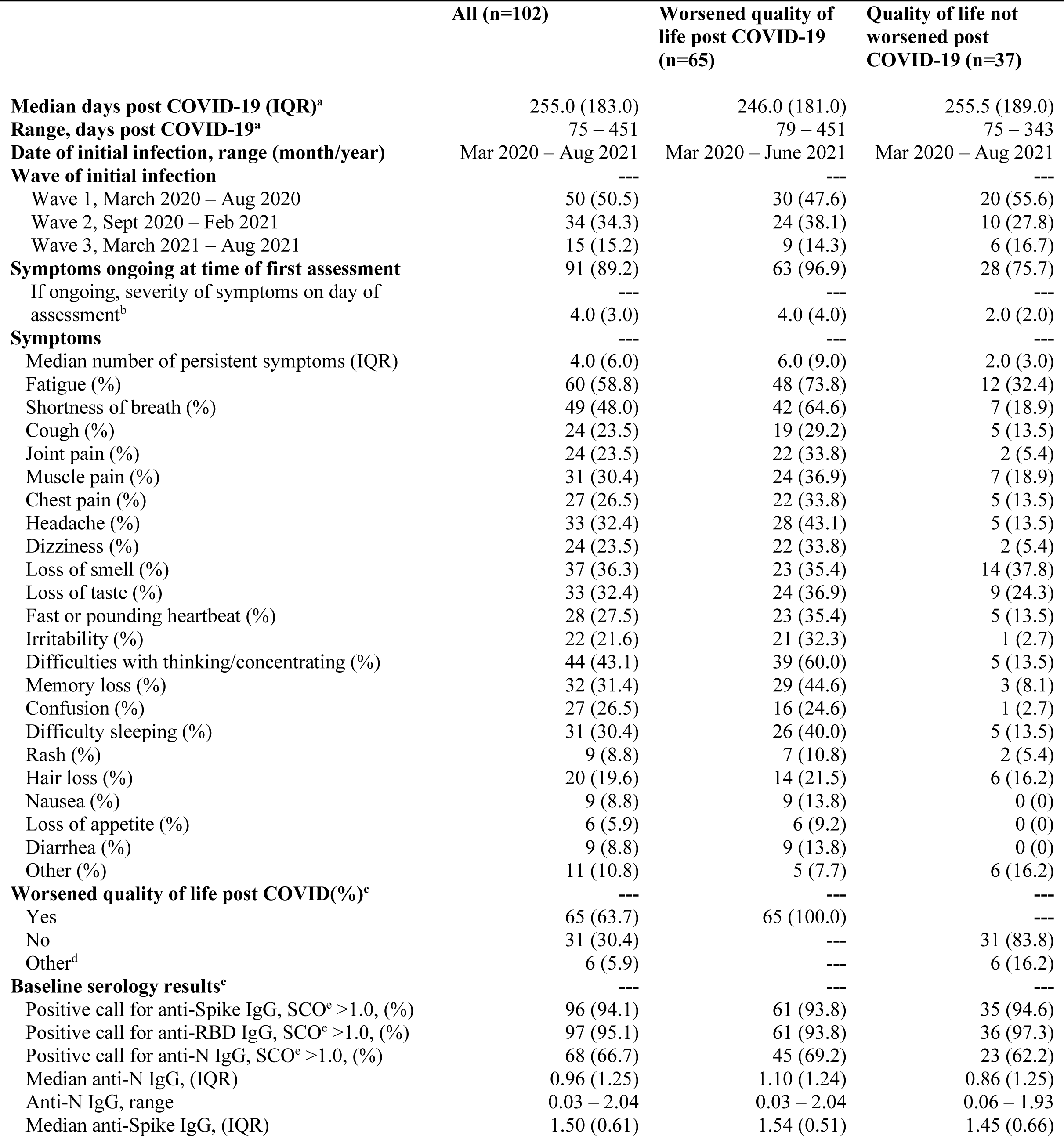
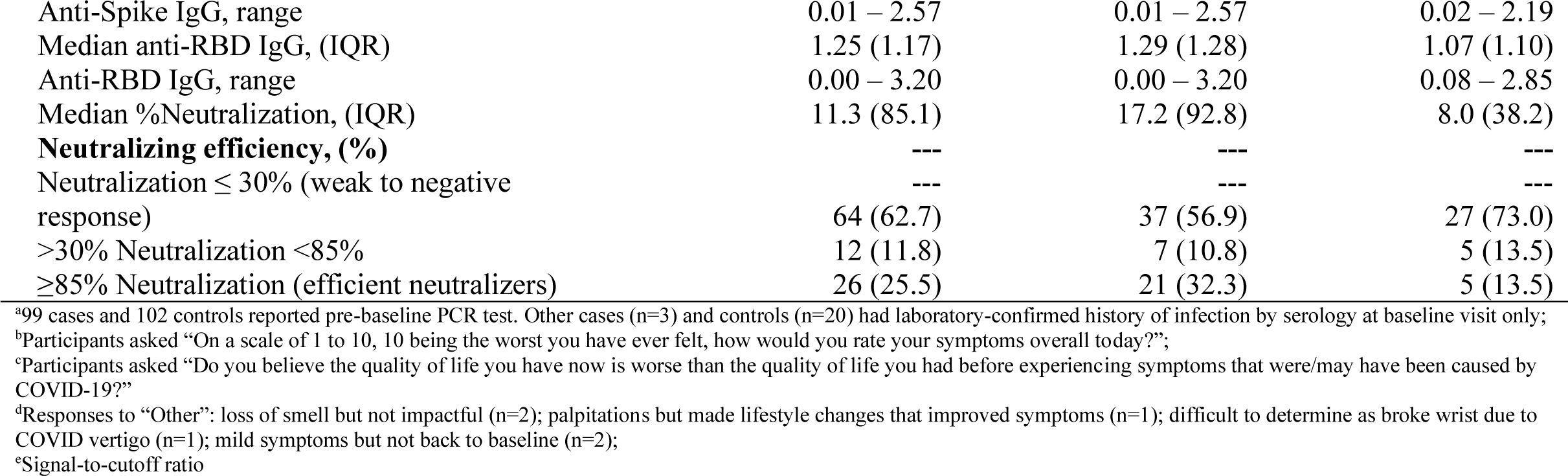
Persisting symptoms (≥12 weeks post COVID-19 onset) and baseline serology of PCC-cases who did (n=65) and did not (n=37) report worsened quality of life.

### Univariate analyses – serological and clinical predictors of PCC

**Table 3** presents crude odds ratios (ORs) with 95% CIs for all variables specified a priori and considered for model inclusion (age, sex, time post-infection (months), asthma requiring medication, conditions/treatments which may suppress the immune system (cancer, HIV, chronic kidney or liver disease, diabetes, organ or bone marrow recipient, other immune deficiency, or receiving treatment that weakens immune system), obesity, smoking, income, allergies, and hospitalization or need for medical attention for COVID-19 symptoms). Unadjusted odds of PCC were 2.3 (95% CI 0.91 – 5.64) given asthma; 2.3 (95% CI 0.73 – 6.99) given diabetes; 1.8 (95% CI 1.04 –3.00) given any allergies; 1.6 (95% CI 0.88 – 2.98) given history of smoking; and 0.7 (95% CI 0.43 – 1.26) given male sex. Not accounting for covariates, participants to have been hospitalized/sought medical attention for COVID-19 symptoms were over four times more likely to have PCC (OR 4.1, 95% CI 2.26 – 7.38). Crude ORs for anti-Spike, anti-RBD, and anti-N titres were non-significant. Unadjusted odds of PCC for efficient neutralizers was 2.3 (95% CI 1.14 – 4.51), as compared to non-efficient neutralizers. Comparing only PCC-cases to report reduced QoL due to symptoms (n=65) with infected-controls (n=122), the crude odds of PCC given efficient neutralization further increased to 3.2 (95% CI 1.53 – 6.62), while negligible increases were observed for other serological predictors (anti-Spike – OR 1.5, 95% CI 0.88 – 2.56; anti-N – OR 1.2, 95% CI 0.69 – 1.92; anti-RBD – OR 1.1, 95% CI 0.74 – 1.58).

**TABLE 3:**
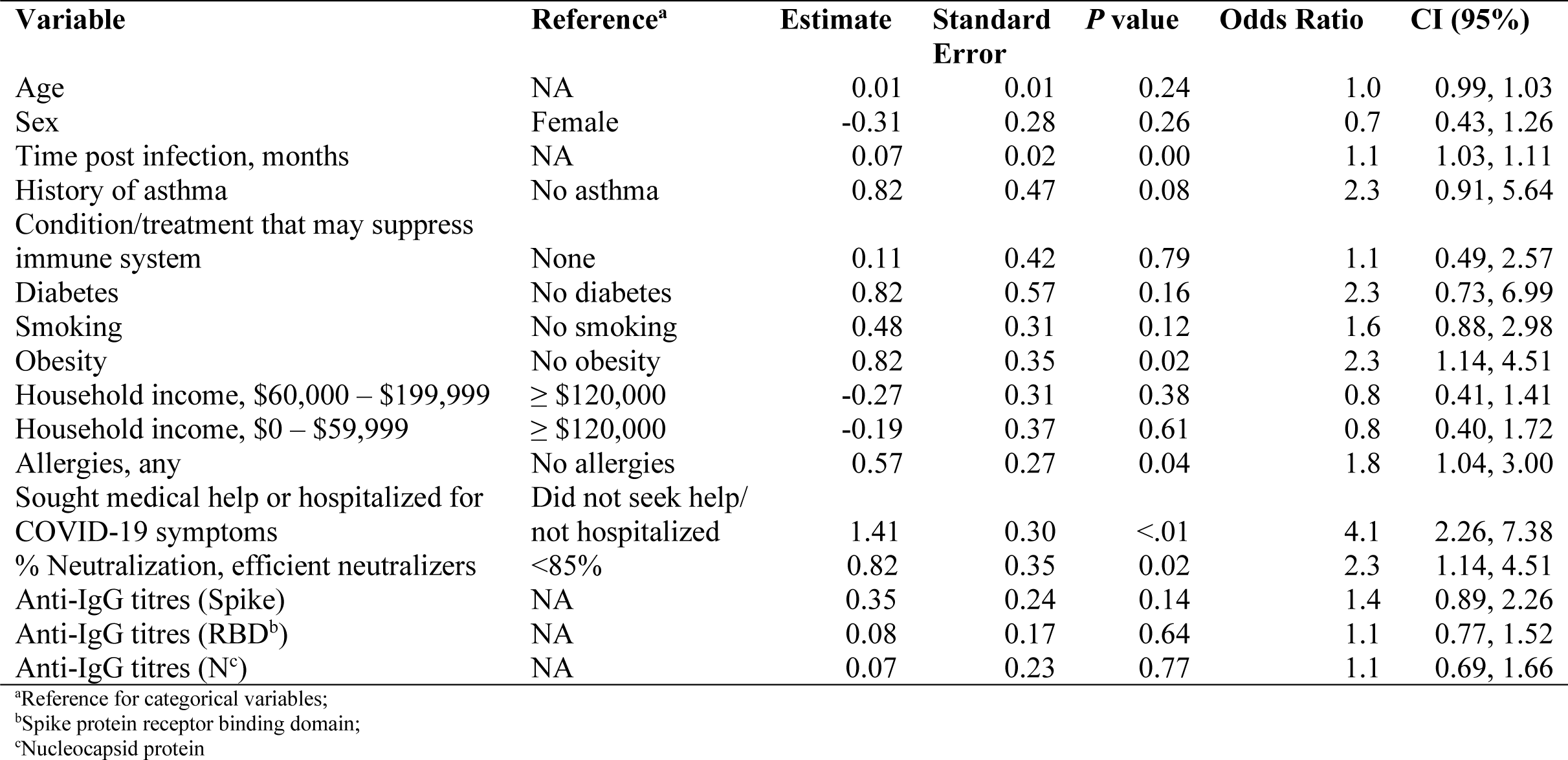
Crude ORs (95%CIs) for Post COVID-19 symptoms: variables considered for analysis prior to modelling.

### Multivariate analyses – serological predictors of PCC accounting for clinical covariates

**Figure 2** compares the effect of IgG titres transformed with restricted cubic splines (k=2, percentiles =10, 50, 90) on odds of PCC in minimally adjusted (covariates – sex and age) and fully adjusted (covariates – sex, age, time since COVID-19 infection, sought medical help/required hospitalization for COVID-19 symptoms, and allergies) models. Using the 10^th^ percentile as the referent and adjusting for age, females tended to have elevated odds of PCC, as compared with males (**Figure 2**). Inter-sex differences in adjusted odds of PCC were reduced in fully adjusted models. Upon testing for pre-specified interactions (each serological predictor and sex; each serological predictor and time post-infection), none were significant.

**FIGURE 2:**
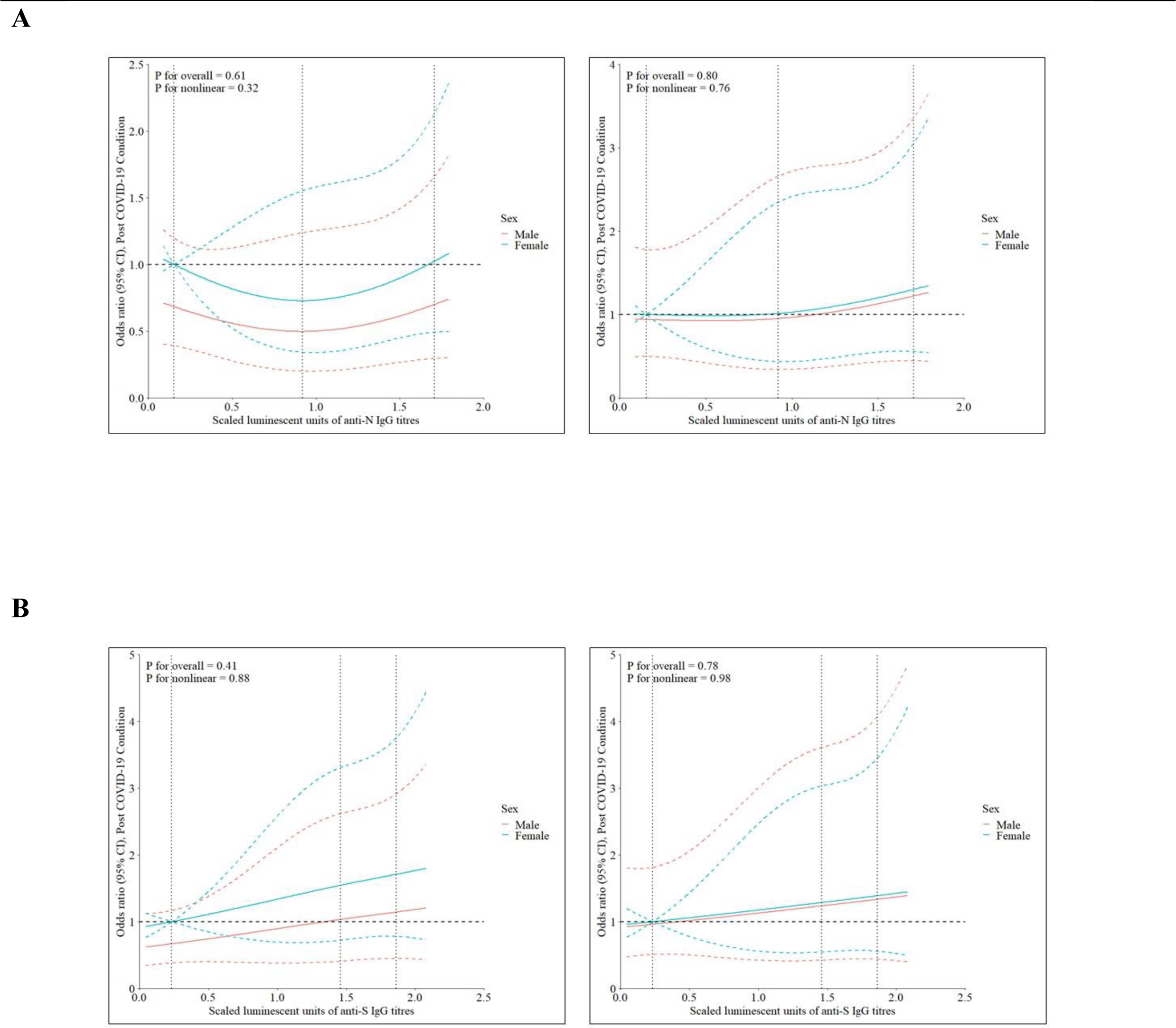
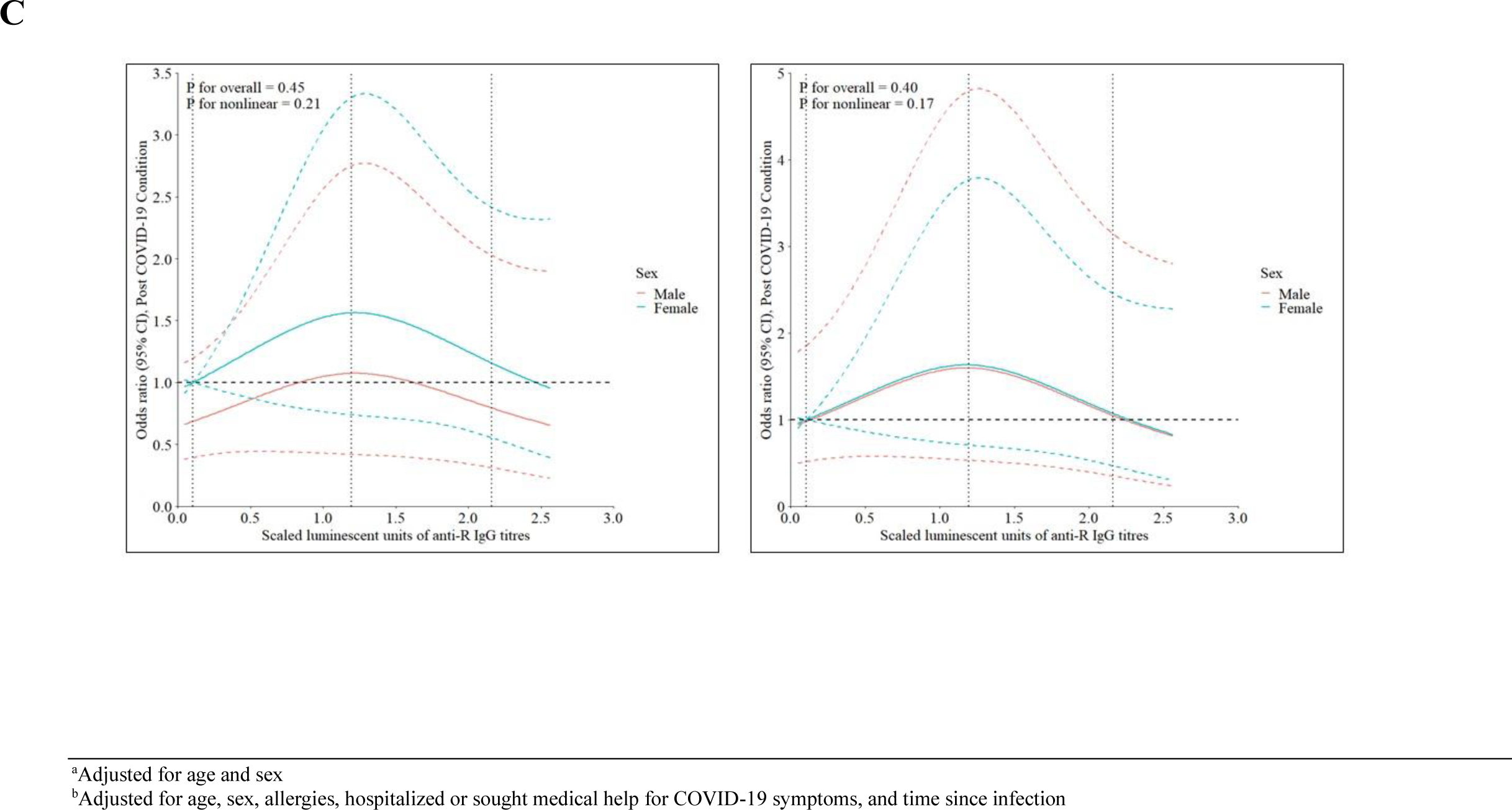
Effect of anti-N (A), anti-Spike (B), and anti RBD (C) IgG titres transformed with restricted cubic splines (k=3, percentiles=10, 50, 90) on odds of Post COVID-19 Condition in minimally^a^ (LEFT) and fully adjusted^b^ (RIGHT) models.

**Tables S3 a-d** present the multivariable model output. Nonlinear relationships between PCC and RCS-transformed predictors were found to be non-significant, with the exception of time post-infection. Area under the curve (AUC) was 0.58 – 0.59 for minimally adjusted models and 0.73 for fully adjusted models (**Figure S1**). Adjusted odds of PCC given changes in IgG titres remained non-significant. Efficient neutralization was a significant predictor of PCC, accounting for age and sex (OR 2.2, 95% CI 1.11 – 4.49), and odds was further increased upon restricting to cases to report reduced quality of life (OR 3.4, 95% CI 1.64 – 7.31). In the fully adjusted model, odds of PCC given neutralizing efficiency ≥85% was 2.0 (95% CI 0.89 – 4.54). Allergies (OR 1.9, 95% CI 1.00 – 3.54), time post-infection (OR 1.9, 95% CI 1.06 – 3.51), and need for hospitalization/medical help for COVID-19 symptoms (OR 3.2, 95% CI 1.61 – 6.24) were associated with PCC in the fully adjusted model (**Table S3d**). Upon removing all cases (10.8%, n=11) and controls (17.2%, n=21) to receive ≥2 COVID-19 vaccines ≥14 days prior to baseline visit, efficient neutralizers had increased odds of PCC in minimally (OR 3.8, 95% CI 1.40 – 10.31) and fully adjusted (OR 2.8, 95% CI 0.91 – 8.31) models. However, these results should be interpreted with caution given the wide confidence intervals, and the result for the fully adjusted model is statistically non-significant. Odds of PCC given neutralization ≥85% also increased upon removal of participants (PCC-cases, n=0; infected-controls, n=5) to receive ≥1 COVID-19 vaccines ≥14 days prior to infection in minimally (OR 2.5, 95% CI 1.22 – 5.19) and fully adjusted (OR 2.2, 95% CI 0.94 – 5.09) models, though the latter remained non-significant.

## Discussion

In our study of a largely non-hospitalized cohort infected with COVID-19 prior to baseline visit, we assessed for relationships between serological markers and PCC, accounting for clinical covariates. We also described the sequelae, quality of life, and health care needs of PCC-cases. Main findings from our study include:

1. Anti-N was a less reliable indicator of past COVID-19 infection than anti-Spike or anti-RBD, among unvaccinated PCC-cases and infected-controls.
2. Among PCC-cases, anti-N, anti-RBD, and anti-Spike IgG titres tended to be higher, as compared to infected-controls. However, associations between PCC and IgG titres remained non-significant in unadjusted and adjusted logistic regression analyses.
3. More PCC-cases were efficient neutralizers (≥85% neutralization) than infected controls. In both unvaccinated and vaccinated subgroups, median neutralization efficiency was somewhat higher among cases than controls.
4. PCC-cases to report worsened quality of life had higher IgG titres and median neutralization efficiency, and a larger proportion of efficient neutralizers, as compared to PCC-cases who did not so report.
5. Clinical covariates associated with PCC and used in multivariate analyses included allergies, time post-infection, and seeking medical help for COVID-19 symptoms.
6. PCC-cases, especially those to report worsened quality of life, were more likely than infected-controls to seek medical help for COVID-19 symptoms, and describe complex and enduring health needs long after initial infection.

### Seropositivity to SARS-COV-2 nucleoprotein, as compared to response elicited by Spike/RBD

Following vaccination for COVID-19, serological response to SARS-CoV-2 nucleoprotein can indicate past infection. However, only 69.2% (n=155) of all participants tested positive for anti-N at baseline. Anti-N seropositivity did not improve upon limiting to participants to have blood drawn 14-90 days post-infection (**Table S2c**). Restricting to non-vaccinated participants (**Table S1a**), more tested positive for anti-Spike and/or anti-RBD than anti-N. Also, PCC-cases were more likely to be seropositive for anti-Spike and/or anti-RBD than infected-controls despite having a significantly higher number of days between infection and first blood draw. We suspect that the sensitivity of SARS-CoV-2 nucleoprotein would have improved given shorter time intervals between infection and serological sampling as anti-N IgG has been found to decay more rapidly than anti-Spike IgG [35–38]. Our findings suggest diminished reliability of anti-N as a marker of past disease as more time elapses between infection and serological testing. Unfortunately, the baseline serological assessment for cases and controls was conducted an average of 4 – 8 months post-infection and we cannot examine for decay at earlier timepoints.

### Associations between Post COVID-19 Condition and IgG titres

Associations between IgG levels and PCC were non-significant in univariate and multivariate analyses. Other studies have found post-infection IgG titres between COVID-19 survivors with and without persistent symptoms to be comparable [39–41], though varying definitions of PCC, time intervals between infection and blood sampling, initial severity of the cohort (e.g., hospitalized vs non-hospitalized), target antigen(s), and other sources of heterogeneity limit comparability of findings. In contrast, García-Abellán *et al.* found lower S1/S2 IgG titres measured at 12 months post-admission to be associated with persistent symptoms (defined as having a score above the third quartile in any items of a self-rated COVID-19 symptom questionnaire at six- and 12-months post-admission) [42]. Blomberg *et al*. reported higher Spike IgG and microneutralizing antibody titres to be associated with both number of persisting symptoms and Chalder Fatigue Scale score assessed six months post-infection [43].

### Neutralization efficiency and Post COVID-19 Condition

Efficient neutralization was significantly associated with PCC, controlling for age and sex. This relationship was further strengthened upon comparing only PCC-cases to report worsened QoL with infected-controls. Neutralizing antibody activity has been found to correlate with initial COVID-19 disease severity [44–46]. Severe acute disease can cause organ damage, immune dysregulation, hypercoagulation, activation of mast cells, and other pathophysiological mechanisms suspected to contribute to PCC [47–48]. Though few in our cohort were hospitalized during acute illness, these results may signify a more robust post-infection response among PCC-cases, especially those with debilitating persisting symptoms. The association between SARS-CoV-2 antibody neutralization and persisting symptoms for three months or longer has been documented previously [49]. However, upon comparing Omicron BA.5 variant and wildtype neutralizing response, Buck *et al.* found only the former to be independently and significantly associated with PCC.

### Clinical predictors and complex medical needs of cases

Clinical covariates found to be strongly associated with PCC included pre COVID-19 allergies and need for hospitalization/medical support. As per **Table 1**, 55.9% of PCC-cases had ≥ 1 allergy, as compared to 41.8% of infected-controls. The two most frequent allergies were reactions to medications (33.3% PCC-cases; 23.0% infected-controls) and pollen (26.5% PCC-cases; 19.9% infected-controls). Cases also reported a wider range of allergies than controls. These findings align with the theory that atypical response to initial infection due to dysfunctional mast cells may manifest as more severe and long-lasting sequelae [50–51]. Allergy status has previously been documented as a potential risk factor for persistent symptoms among adults and children [51–54]. However, findings are especially limited among adult cohorts and more research on allergic phenotypes is required [53].

During analysis, we noted that need for medical attention may not exclusively refer to supports sought in the acute phase of illness. A few participants (**Table 4**) reported seeking medical attention for chronic symptoms post-infection (e.g., one case described seeking help at the Post COVID-19 care clinic). Unfortunately, it was often not possible to discern from our cohort when help was sought post disease onset. While this data issue limits the value of this variable as a proxy for acute illness severity, our findings that more PCC-cases required medical attention (50.0%), as well as different types of medical attention (2-5 types required by 23.0%), as compared with infected-controls, are important in the context of understanding the complex health needs of people with PCC. We also found that cases who reported worsened QoL post COVID-19 were more likely to require medical support (61.5%), as compared with cases who did not report worsened QoL. The most common sources of medical support sought by cases were 1) family doctor/primary care provider (29.4% for all cases; 43.1% for cases with worsened QoL); 2) public health testing centre (20.6% for all cases; 24.6% for cases with worsened QoL); and 3) emergency department (20.6% for all cases; 27.7% for cases with worsened QoL), **Table 4**. Findings from K10 and IES surveys also suggest a higher burden of mental health needs among PCC-cases (**Table 1**), and **Table 2** describes high diversity, severity, and longevity of PCC symptoms among cases. At time of first assessment, 89.2% of cases (n=91) reported symptoms persisting long after initial disease (median 255 days, IQR 183). Given ongoing labour shortages in the healthcare sector, many people with PCC may not gain timely access to care. Our findings support the need for ample staff and resources to respond to prolonged, recurrent, and diverse needs across multiple health domains. For example, more Post COVID-19 care clinics may improve the well-being, function, and quality of life of this population while reducing burden on the mainstream health system [55–61].

**TABLE 4:** Type of medical attention sought for COVID-19 symptoms.

|  | Cases (n=102)<sup>a</sup> | Cases with worsened quality of life (n=65) | Controls (n=122)<sup>b</sup> |
| --- | --- | --- | --- |
| **Type of medical attention sought, %** | 51 (50.0) | 40 (61.5) | 24 (19.7) |
| Family doctor/primary care provider | 30 (29.4) | 28 (43.1) | 5 (4.1) |
| Occupational Health | 7 (6.9) | 6 (9.2) | 2 (1.6) |
| Public Health Testing Centre | 21 (20.6) | 16 (24.6) | 9 (7.4) |
| Walk-in or Urgent Care Clinic | 3 (2.9) | 3 (4.6) | 1 (0.8) |
| Emergency Department | 21 (20.6) | 18 (27.7) | 8 (6.6) |
| Telehealth | 6 (5.9) | 5 (7.7) | 1 (0.8) |
| Other<sup>a</sup> | 7 (6.9) | 6 (9.2) | 2 (1.6) |
| **Number of types of medical attention sought, %** | --- | --- | --- |
| 1 | 28 (27.5) | 21 (32.3) | 20 (16.4) |
| 2 | 7 (6.9) | 7 (10.8) | 4 (3.3) |
| 3 | 11 (10.8) | 9 (13.8) | 0 (0) |
| 4 | 4 (3.9) | 4 (6.2) | 0 (0) |
| 5 | 1 (1.0) | 1 (1.5) | 0 (0) |

<sup>a</sup>“Other” for Cases includes Naturopath = 1; Naturopath and physiotherapist=2; Ottawa Public Health=1; Physiotherapist=1; Physiotherapist, massage therapist, and respiratory therapist=1; Post COVID care clinic=1;“Other” for Controls includes Oral Surgeon - bacteria on my tongue=1; Blood tests EKG performed related to chest pains after first dose of vaccine=1

### Limitations

Several limitations may have influenced findings. First, most of our participants were not hospitalized for COVID-19. Given that people with more severe disease tend to elicit higher antibody titres [62–63], results may have varied if more of our cohort required hospitalization during acute illness. Second, participants had limited diversity in terms of age, race, employment, pre COVID-19 comorbidities, and income status. Most reported high household income, were well-educated and employed pre COVID-19, and generally healthy. A study sample more representative of the total population at risk of PCC may have generated different findings. Third, as most clinical data was self-reported through electronic questionnaires, there is risk of response bias. Fourth, results from subgroup analyses of varying post-infection intervals (**Tables S2a-c**) were limited by smaller sample sizes, and wave of infection was a potentially confounding factor. For example, only one case and two controls infected March 2020 - August 2020 had blood drawn 14-90 days post-infection (**Table S2c**). Fifth, study sample size also limited opportunity to assess for differences in serological response as a function of PCC subtypes. There is poor consensus on how subtypes of PCC character and severity should be defined [64]. We used self-reported quality of life due to persistent symptoms as a proxy of PCC severity. Other studies have found that certain PCC symptoms/clusters correlate with stronger or weaker serological response post COVID-19. For example, Molnar *et al.* found serum levels of anti-Spike IgG and anti-N IgG to be significantly lower in patients with severe fatigue post COVID-19, as compared to patients with non-severe fatigue [65], while Su *et al.* reported high anti-N IgG in cases of neurological PCC [66]. Sixth, as only five controls and no cases received ≥1 COVID-19 vaccines ≥14 days prior infection, this study afforded limited opportunity to examine the protective capacity of hybrid immunity [67]. Seventh, in lieu of established cut-offs for neutralization efficiency, these were derived by study team members to develop the in-house snELISA used in SSO. Lastly, the availability of IgG antibody subtypes would have permitted more detailed analyses.

### Next steps

Given highly mixed results in the literature, we are undertaking a robust review on post-acute serological predictors of PCC: https://www.crd.york.ac.uk/PROSPERO/display_record.php?RecordID=402978. This strategy will allow us to examine trends in serological markers and associations with persistent sequelae among multiple studies with varying cohort characteristics and procedures to collect and analyze post-acute findings. We will investigate and report on different sources of heterogeneity which may influence trends. We will also attempt to integrate review estimates in future analyses of SSO study data through use of Bayesian logistic regression. Given our limited sample size and distribution, encompassing prior information from the literature will facilitate more detailed and diverse analyses of serological predictors, accounting for clinical covariates. Finally, assessment of IgG subtypes will be pursued once available.

### Conclusion

In summary, we found associations between Post COVID-19 Condition (PCC) and anti-N, anti-Spike, and anti-RBD IgG titres to be non-significant. However, as compared to infected-controls, PCC-cases had significantly higher neutralization efficiency, especially those to report deteriorated quality of life. Future investigations of IgG subtypes and PCC symptom clusters may elucidate new findings. Comparison with other studies is hampered by gross heterogeneity in cohort characteristics, definitions of PCC, and laboratory procedures. Standardized reporting of PCC and serological results would advance current efforts to collate and analyze inter-study findings. Finally, cases with PCC reported complex, ongoing sequelae a median of 255 days (183 IQR) post COVID-19, which underscores the potential need for health supports and services long after infection.

## Acknowledgements

The authors acknowledge all investigators and partner organizations contributing to the project: CIs: Steffany Bennett, Pranesh Chakraborty, Miroslava Culf-Cuperlovic, Yves Durocher, Jennifer Quaid, Mary Simmerling. Partner organizations: National Research Council (NRC) of Canada, the Ottawa Hospital Research Institute, the University of Ottawa, and CoVaRR-Net (Coronavirus Variants Rapid Response Network). COVID-19 reagents (viral antigens, ACE2-biotin and anti-human IgG-HRP fusion) were generously provided by Dr Yves Durocher at the NRC Montreal. UOttawa Serology and Diagnostics High Throughput Facility is supported by Danielle Dewar-Darch, Justino Hernandez Soto, and Abishek Xavier.

## Author contributions

M-AL, EC, and JL drafted the manuscript. JL, RS, EC, RB, CAB, CLC, AMC, MM, and M-AL were involved in the conception and design of the Stop the Spread Ottawa study. EC performed analyses. CG and SH provided statistical support. YG, CA, PM, MP, and LR significantly contributed to serological assay development, implementation, planning, and analyses. AMC and M-AL coordinated all laboratory processing of cohort biological specimens. M-AL is responsible for the overall content as the guarantor. All authors critically reviewed and approved the final manuscript.

## Funding

EC is supported by the AI4PH Scholarship Program, funded by CIHR (Canadian Institutes of Health Research - Instituts de recherche en santé du Canada). YG is supported by a Charles Best and Frederick Banting (CGS-Doctoral award) from CIHR (476885). The Stop the Spread Ottawa study is funded by CIHR (424425), the COVID-19 Immunity Task Force (CITF), and the University of Ottawa. The study extension is funded by the Coronavirus Variants Rapid Response Network (CoVaRR-Net) (156941) (https://covarrnet.ca/investigating-long-term-variables-to-sars-cov-2-infection-and-vaccine-immunity/). CoVaRR-Net is funded by an operating grant from CIHR (FRN# 175622). We also acknowledge in-kind support from the NRC’s Pandemic Response Challenge Program.

## Competing interests

None declared

## Patient and public involvement

Patients and/or the public were involved in the design, or conduct, or reporting, or dissemination plans of this research.

## Ethics approval

Ethics approval was obtained from the The Ottawa Health Science Network Research Ethics Board (Protocol ID Number: 20200481-01H), and access to the data sets was granted by relevant data custodians.

## Data availability statement

Direct access to the data and analytical files is not permitted without the expressed permission of the approving human research ethics committees and data custodians. Researchers interested in collaboration should contact the corresponding authors.

## Notes

### Competing Interest Statement

The authors have declared no competing interest.

### Funding Statement

EC is supported by the AI4PH Scholarship Program, funded by CIHR. YG is supported by a Charles Best and Frederick Banting (CGS-Doctoral award) from CIHR (476885). The Stop the Spread Ottawa study is funded by the Canadian Institutes of Health Research (CIHR) (424425), the COVID-19 Immunity Task Force (CITF), and the University of Ottawa. The study extension is funded by the Coronavirus Variants Rapid Response Network (CoVaRR-Net) (156941). CoVaRR-Net is funded by an operating grant from CIHR (FRN 175622). We also acknowledge in-kind support from the NRC Pandemic Response Challenge Program.

## References

1. CDC. COVID-19 and Your Health. Centers for Disease Control and Prevention. Published December 16, 2022. https://www.cdc.gov/coronavirus/2019-ncov/long-term-effects/index.html

2. Canada PHA of. Post-COVID-19 condition (long COVID). Published August 20, 2021. https://www.canada.ca/en/public-health/services/diseases/2019-novel-coronavirus-infection/symptoms/post-covid-19-condition.htmlday

3. Long Covid Impact on Adult Americans: Early Indicators Estimating Prevalence and Cost. Solve Long COVID Initiative, April 5, 2022. https://solvecfs.org/wp-content/uploads/2022/04/Long_Covid_Impact_Paper.pdf

4. Government of Canada SC. Canadian COVID-19 Antibody and Health Survey (CCAHS). Published October 13, 2022. https://www23.statcan.gc.ca/imdb/p2SV.pl?Function=getSurvey&SDDS=5339

5. Davis HE, McCorkell L, Vogel JM, Topol EJ. Long COVID: major findings, mechanisms and recommendations. Nat Rev Microbiol. 2023;21(3):133–146. doi:10.1038/s41579-022-00846-2

6. Understanding the Post COVID-19 Condition (Long COVID) in Adults and the Expected Burden for Ontario. Ontario COVID-19 Science Advisory Table. doi:10.47326/ocsat.2022.03.65.1.0

7. Castanares-Zapatero D, Chalon P, Kohn L, et al. Pathophysiology and mechanism of long COVID: a comprehensive review. Annals of medicine. 2022;54(1). doi:10.1080/07853890.2022.2076901

8. Perlis RH, Santillana M, Ognyanova K, et al. Prevalence and Correlates of Long COVID Symptoms Among US Adults. JAMA Netw Open. 2022;5(10):e2238804–e2238804. doi:10.1001/jamanetworkopen.2022.38804

9. Post-COVID-19 Condition in Canada: What we know, what we don’t know, and a framework for action. Published May 17, 2023. https://science.gc.ca/site/science/en/office-chief-science-advisor/initiatives-covid-19/post-covid-19-condition-canada-what-we-know-what-we-dont-know-and-framework-action

10. Chen C, Haupert SR, Zimmermann L, et al. Global Prevalence of Post COVID-19 Condition or Long COVID: A Meta-Analysis and Systematic Review. J Infect Dis. Published online April 16, 2022:jiac136. doi:10.1093/infdis/jiac136

11. Azzolini E, Levi R, Sarti R, et al. Association Between BNT162b2 Vaccination and Long COVID After Infections Not Requiring Hospitalization in Health Care Workers. JAMA. 2022;328(7):676–678. doi:10.1001/jama.2022.11691

12. Fernández-de-las-Peñas C, Notarte KI, Peligro PJ, et al. Long-COVID Symptoms in Individuals Infected with Different SARS-CoV-2 Variants of Concern: A Systematic Review of the Literature. Viruses. 2022;14(12). doi:10.3390/v14122629

13. Byambasuren O, Stehlik P, Clark J, et al. Effect of covid-19 vaccination on long covid: systematic review. BMJ Medicine. 2023;2(1). doi:10.1136/bmjmed-2022-000385

14. Notarte KI, Catahay JA, Velasco JV, et al. Impact of COVID-19 vaccination on the risk of developing long-COVID and on existing long-COVID symptoms: A systematic review. eClinicalMedicine. 2022;53. doi:10.1016/j.eclinm.2022.101624

15. Munblit D, Nicholson TR, Needham DM, et al. Studying the post-COVID-19 condition: research challenges, strategies, and importance of Core Outcome Set development. BMC Med. 2022;20(1):1–13. doi:10.1186/s12916-021-02222-y

16. Cervia C, Zurbuchen Y, Taeschler P, et al. Immunoglobulin signature predicts risk of post-acute COVID-19 syndrome. Nat Commun. 2022;13(1):1–12. doi:10.1038/s41467-021-27797-1

17. Knight DRT, Munipalli B, Logvinov II, et al. Perception, Prevalence, and Prediction of Severe Infection and Post-acute Sequelae of COVID-19. The American Journal of the Medical Sciences. 2022;363(4):295–304. doi:10.1016/j.amjms.2022.01.002

18. Fabbri A, Voza A, Riccardi A, Vanni S, De Iaco F. Unfavorable Outcome and Long-Term Sequelae in Cases with Severe COVID-19. Viruses. 2023;15(2):485. doi:10.3390/v15020485

19. Tsampasian V, Elghazaly H, Chattopadhyay R, et al. Risk Factors Associated With Post−COVID-19 Condition: A Systematic Review and Meta-analysis. JAMA Intern Med. Published online March 23, 2023. doi:10.1001/jamainternmed.2023.0750

20. Jia X, Cao S, Lee AS, et al. Anti-nucleocapsid antibody levels and pulmonary comorbid conditions are linked to post–COVID-19 syndrome. JCI Insight. 2022;7(13). doi:10.1172/jci.insight.156713

21. Krishna BA, Lim EY, Mactavous L, et al. Evidence of previous SARS-CoV-2 infection in seronegative patients with long COVID. eBioMedicine. 2022;81:104129. doi:10.1016/j.ebiom.2022.104129

22. García-Abellán J, Padilla S, Fernández-González M, et al. Antibody Response to SARS-CoV-2 is Associated with Long-term Clinical Outcome in Patients with COVID-19: a Longitudinal Study. J Clin Immunol. 2021;41(7):1490–1501. doi:10.1007/s10875-021-01083-7

23. Gaebler C, Wang Z, Lorenzi JCC, et al. Evolution of antibody immunity to SARS-CoV-2. Nature. 2021;591(7851):639–644. doi:10.1038/s41586-021-03207-w

24. Blomberg B, Mohn KGI, Brokstad KA, et al. Long COVID in a prospective cohort of home-isolated patients. Nat Med. 2021;27(9):1607–1613. doi:10.1038/s41591-021-01433-3

25. García-Abellán J, Fernández M, Padilla S, et al. Immunologic phenotype of patients with long-COVID syndrome of 1-year duration. Front Immunol. 2022;13. doi:10.3389/fimmu.2022.920627

26. Gerhards C, Thiaucourt M, Kittel M, et al. Longitudinal assessment of anti-SARS-CoV-2 antibody dynamics and clinical features following convalescence from a COVID-19 infection. International Journal of Infectious Diseases. 2021;107:221–227. doi:10.1016/j.ijid.2021.04.080

27. Lier J, Stoll K, Obrig H, et al. Neuropsychiatric phenotype of post COVID-19 syndrome in non-hospitalized patients. Front Neurol. 2022;13. doi:10.3389/fneur.2022.988359

28. Augustin M, Schommers P, Stecher M, et al. Post-COVID syndrome in non-hospitalised patients with COVID-19: a longitudinal prospective cohort study. The Lancet Regional Health -Europe. 2021;6:100122. doi:10.1016/j.lanepe.2021.100122

29. Durstenfeld MS, Peluso MJ, Kelly JD, et al. Role of antibodies, inflammatory markers, and echocardiographic findings in postacute cardiopulmonary symptoms after SARS-CoV-2 infection. JCI Insight. 2022;7(10). doi:10.1172/jci.insight.157053

30. Collins E, Galipeau Y, Arnold C, et al. Cohort profile: Stop the Spread Ottawa (SSO)—a community-based prospective cohort study on antibody responses, antibody neutralisation efficiency and cellular immunity to SARS-CoV-2 infection and vaccination. BMJ Open. 2022;12(9):e062187. doi:10.1136/bmjopen-2022-062187

31. Colwill K, Galipeau Y, Stuible M, et al. A scalable serology solution for profiling humoral immune responses to SARS-CoV-2 infection and vaccination. Clinical & Translational Immunology. 2022;11(3):e1380. doi:10.1002/cti2.1380

32. Kessler RC, Andrews G, Colpe LJ, et al. Short screening scales to monitor population prevalences and trends in non-specific psychological distress. Psychological Medicine. 2002;32(6):959–976. doi:10.1017/S0033291702006074

33. Horowitz M, Wilner N, Alvarez W, et al. Impact of Event Scale: a measure of subjective stress. Psychosomatic medicine. 1979;41(3). doi:10.1097/00006842-197905000-00004

34. Harrell, FE. Regression modeling strategies. Second edition, Springer, 2015.

35. Elslande JV, Oyaert M, Lorent N, et al. Lower persistence of anti-nucleocapsid compared to anti-spike antibodies up to one year after SARS-CoV-2 infection. Diagnostic Microbiology and Infectious Disease. 2022;103(1):115659. doi:10.1016/j.diagmicrobio.2022.115659

36. Van den Hoogen LL, Smits G, van Hagen CCE, et al. Seropositivity to Nucleoprotein to detect mild and asymptomatic SARS-CoV-2 infections: A complementary tool to detect breakthrough infections after COVID-19 vaccination? Vaccine. 2022;40(15):2251–2257. doi:10.1016/j.vaccine.2022.03.009

37. Krutikov M, Palmer T, Tut G, et al. Prevalence and duration of detectable SARS-CoV-2 nucleocapsid antibodies in staff and residents of long-term care facilities over the first year of the pandemic (VIVALDI study): prospective cohort study in England. The Lancet Healthy Longevity. 2022;3(1):e13–e21. doi:10.1016/S2666-7568(21)00282-8

38. Whitaker HJ, Gower C, Otter AD, et al. Nucleocapsid antibody positivity as a marker of past SARS-CoV-2 infection in population serosurveillance studies: impact of variant, vaccination, and choice of assay cut-off. medRxiv. Published online October 26, 2021:2021.10.25.21264964. doi:10.1101/2021.10.25.21264964

39. Seeßle J, Waterboer T, Hippchen T, et al. Persistent Symptoms in Adult Patients 1 Year After Coronavirus Disease 2019 (COVID-19): A Prospective Cohort Study. Clin Infect Dis. 2022;74(7):1191–1198. doi:10.1093/cid/ciab611

40. Peluso MJ, Lu S, Tang AF, et al. Markers of Immune Activation and Inflammation in Individuals With Postacute Sequelae of Severe Acute Respiratory Syndrome Coronavirus 2 Infection. J Infect Dis. 2021;224(11):1839–1848. doi:10.1093/infdis/jiab490

41. Ozonoff A, Schaenman J, Jayavelu ND, et al. Phenotypes of disease severity in a cohort of hospitalized COVID-19 patients: Results from the IMPACC study. eBioMedicine. 2022;83. doi:10.1016/j.ebiom.2022.104208

42. García-Abellán J, Fernández M, Padilla S, et al. Immunologic phenotype of patients with long-COVID syndrome of 1-year duration. Front Immunol. 2022;13:920627. doi:10.3389/fimmu.2022.920627

43. Blomberg B, Mohn KGI, Brokstad KA, et al. Long COVID in a prospective cohort of home-isolated patients. Nat Med. 2021;27(9):1607–1613. doi:10.1038/s41591-021-01433-3

44. Sancilio A, Schrock JM, Demonbreun AR, et al. COVID-19 symptom severity predicts neutralizing antibody activity in a community-based serological study. Sci Rep. 2022;12(1):1–7. doi:10.1038/s41598-022-15791-6

45. Garcia-Beltran WF, Lam EC, Astudillo MG, et al. COVID-19-neutralizing antibodies predict disease severity and survival. Cell. 2021;184(2):476–488.e11. doi:10.1016/j.cell.2020.12.015

46. Liu L, To KKW, Chan KH, et al. High neutralizing antibody titer in intensive care unit patients with COVID-19. Emerging Microbes & Infections. Published online July 20, 2020. https://www.tandfonline.com/doi/abs/10.1080/22221751.2020.1791738

47. Castanares-Zapatero D, Chalon P, Kohn L, et al. Pathophysiology and mechanism of long COVID: a comprehensive review. Annals of medicine. 2022;54(1). doi:10.1080/07853890.2022.2076901

48. Batiha GES, Al-kuraishy HM, Al-Gareeb AI, Welson NN. Pathophysiology of Post-COVID syndromes: a new perspective. Virol J. 2022;19(1):1–20. doi:10.1186/s12985-022-01891-2

49. Buck AM, Deitchman AN, Takahashi S, et al. The Breadth of the Neutralizing Antibody Response to Original SARS-CoV-2 Infection is Linked to the Presence of Long COVID Symptoms. medRxiv. 2023:2023.03.30.23287923. doi:10.1101/2023.03.30.23287923

50. Afrin LB, Weinstock LB, Molderings GJ. Covid-19 hyperinflammation and post-Covid-19 illness may be rooted in mast cell activation syndrome. International Journal of Infectious Diseases. 2020;100:327–332. doi:10.1016/j.ijid.2020.09.016

51. Osmanov IM, Spiridonova E, Bobkova P, et al. Risk factors for post-COVID-19 condition in previously hospitalised children using the ISARIC Global follow-up protocol: a prospective cohort study. European Respiratory Journal. 2022;59(2). doi:10.1183/13993003.01341-2021

52. Kostev K, Smith L, Koyanagi A, Konrad M, Jacob L. Post-COVID-19 conditions in children and adolescents diagnosed with COVID-19. Pediatr Res. Published online May 14, 2022:1–6. doi:10.1038/s41390-022-02111-x

53. Jacobs ET, Catalfamo CJ, Columbo PM, et al. Pre-existing conditions associated with post-acute sequelae of COVID-19. Journal of Autoimmunity. 2023;135:102991. doi:10.1016/j.jaut.2022.102991

54. Bell ML, Catalfamo CJ, Farland LV, et al. Post-acute sequelae of COVID-19 in a non-hospitalized cohort: Results from the Arizona CoVHORT. PLOS ONE. 2021;16(8):e0254347. doi:10.1371/journal.pone.0254347

55. Santhosh L, Block B, Kim SY, et al. Rapid design and implementation of Post-COVID-19 Clinics. Chest. 2021;160(2):671. doi:10.1016/j.chest.2021.03.044

56. Ibrahim Mohamed HM, Atya M sayed, Ahmed GH, Abdelmohsen SA, Sobhy KM, Hussein AM. Telehealth program: Effect of Physiotherapy Intervention on Dyspnea, Fatigue and Functional Status of Post COVID-19 Syndrome patients. Assiut Scientific Nursing Journal. 2022;10(32):265–280. doi:10.21608/asnj.2022.167006.1437

57. Houben S, Bonnechère B. The Impact of COVID-19 Infection on Cognitive Function and the Implication for Rehabilitation: A Systematic Review and Meta-Analysis. International Journal of Environmental Research and Public Health. 2022;19(13):7748. doi:10.3390/ijerph19137748

58. Garg A, Subramain M, Barlow PB, et al. Patient Experiences with a Tertiary Care Post-COVID-19 Clinic. Journal of Patient Experience. Published online January 17, 2023. doi:10.1177/23743735231151539

59. Nowakowski S, Kokonda M, Sultana R, et al. Association between Sleep Quality and Mental Health among Patients at a Post-COVID-19 Recovery Clinic. Brain Sciences. 2022;12(5):586. doi:10.3390/brainsci12050586

60. Levan S, Mourad M, Block B, Shah R, Santhosh L. Impact of a Multidisciplinary Post-COVID-19 Clinic on Hospital Admissions and ED Visits. Chest. Published online January 2023:S0012369223000090. doi:10.1016/j.chest.2022.12.031

61. Kotb N, Barreto L, Janaudis-Ferreira T. Post-exertional malaise in pulmonary rehabilitation after COVID-19: Are we not giving enough attention? Canadian Journal of Respiratory, Critical Care, and Sleep Medicine. 2023;7(2):93–118. doi:10.1080/24745332.2022.2150722

62. Park JH, Cha MJ, Choi H, et al. Relationship between SARS-CoV-2 antibody titer and the severity of COVID-19. Journal of Microbiology, Immunology and Infection. 2022;55(6):1094–1100. doi:10.1016/j.jmii.2022.04.005

63. Yan X, Chen G, Jin Z, et al. Anti-SARS-CoV-2 IgG levels in relation to disease severity of COVID-19. Journal of Medical Virology. 2022;94(1):380–383. doi:10.1002/jmv.27274

64. Basharat S, Chao YS, McGill SC. Subtypes of Post–COVID-19 Condition: A Review of the Emerging Evidence. cjht. 2022;2(12). doi:10.51731/cjht.2022.516

65. Molnar T, Varnai R, Schranz D, et al. Severe fatigue and memory impairment are associated with lower serum level of anti-SARS-CoV-2 antibodies in patients with post-COVID symptoms. Journal of Clinical Medicine. 2021;10(19):4337. doi:10.3390/jcm10194337

66. Su Y, Yuan D, Chen DG, et al. Multiple early factors anticipate post-acute COVID-19 sequelae. Cell. 2022;185(5):881–895.e20. doi:10.1016/j.cell.2022.01.014

67. Bobrovitz N, Ware H, Ma X, et al. Protective effectiveness of previous SARS-CoV-2 infection and hybrid immunity against the omicron variant and severe disease: a systematic review and meta-regression. The Lancet Infectious Diseases. 2023;23(5):556–567. doi:10.1016/S1473-3099(22)00801-5

