## Supplemental Files for "CLINICAL AND SEROLOGICAL PREDICTORS OF POST COVID-19 CONDITION – FINDINGS FROM A CANADIAN PROSPECTIVE COHORT STUDY"

| **Table S1a: Serological findings of PCC-cases and infected-controls with baseline assessment ≥14 days prior to receiving ≥1** **COVID-19 vaccines (n=171)** | | | |
| --- | --- | --- | --- |
|  | **Cases (n=77)^a^** | **Controls (n=94)^b^** | ***P* value^c^** |
| **Days between COVID-19 infection and baseline serology**  Median (IQR)  Range | ---  205 (163.0)  12 – 414 | ---  84 (152.0)  13 – 384 | ---  <0.01  --- |
| **Baseline serology results**  Positive call for anti-Spike IgG (SCO >1, %)^d^ | ---  71 (92.2) | ---  81 (86.2) | ---  0.23 |
| Positive call for anti-RBD IgG (SCO >1, %)^d^ | 72 (93.5) | 75 (79.8) | 0.01 |
| Positive call for anti-N IgG (SCO >1, %)^d^ | 50 (64.9) | 66 (70.2) | 0.51 |
| Median anti-N IgG, (IQR) | 0.89 (1.21) | 0.90 (1.06) | 0.89 |
| Anti-N IgG, range | 0.03 – 2.04 | 0.07 – 2.01 | --- |
| Median anti-Spike IgG, (IQR) | 1.45 (0.79) | 1.29 (0.86) | 0.14 |
| Anti-Spike IgG, range | 0.01 – 2.57 | 0.01 – 2.58 | --- |
| Median anti-RBD IgG, (IQR) | 0.87 (1.24) | 0.89 (1.37) | 0.54 |
| Anti-RBD IgG, range | 0.00 – 3.20 | 0.00 – 2.80 | --- |
| Median %Neutralization, (IQR) | 7.17 (20.35) | 3.62 (17.40) | 0.45 |
| %Neutralization, range | 0.00 – 99.09 | 0.00 – 97.78 | --- |
| **Neutralizing efficiency, (%)** | --- | --- | --- |
| Neutralization ≤ 30% (weak to negative response) | 63 (81.8) | 76 (80.9) | 0.39 |
| >30% Neutralization <85% | 9 (11.7) | 16 (17.0) | REF |
| ≥85% Neutralization (efficient neutralizers) | 5 (6.5) | 2 (2.1) | 0.11 |
| ^a^ Number missing for variables, Cases: PCR test date 3, Vaccination dates 1;  ^b^ Number missing for variables, Controls: PCR test date 13;  ^c^ *P* value <0.05: chi-squared and Fisher’s exact tests used for categorical variables; Wilcoxon Rank Sum test used for continuous variables;  ^d^ Signal-to-cutoff ratio | | | |

| **Table S1b: Serological findings of PCC-cases and infected-controls to receive ≥1** **COVID-19** **vaccines ≥14 days prior to baseline visit (n=52)** | | | |
| --- | --- | --- | --- |
|  | **Cases (n=24)^a^** | **Controls (n=28)^b^** | ***P* value^c^** |
| **Number of vaccines received ≥14 days prior to baseline visit**  1 dose  2 doses | ---  13 (54.2)  11 (45.8) | ---  7 (25.0)  21 (75.0) | *---*  0.05  0.05 |
| **Days between first COVID-19 vaccine and baseline serology**  Median (IQR)  Range  **Days between second COVID-19 vaccine and baseline serology**  Median (IQR)  Range | ---  65 (59.5)  14 – 206  ---  62 (42.0)  21 – 121 | ---  102 (76.0)  20 – 208  ---  63 (29.0)  19 – 109 | ---  0.08  ---  ---  1.00  --- |
| **Days between COVID-19 infection and baseline serology**  Median (IQR)  Range | ---  162 (112.5)  76 – 414 | ---  147 (44.0)  14 – 333 | ---  0.12  --- |
| **Baseline serology results**  Positive call for anti-Spike IgG (SCO >1, %)^d^ | ---  24 (100.0) | ---  28 (100.0) | ---  1.00 |
| Positive call for anti-RBD IgG (SCO >1, %)^d^ | 24 (100.0) | 28 (100.0) | 1.00 |
| Positive call for anti-N IgG (SCO >1, %)^d^ | 17 (70.8) | 21 (75.0) | 0.76 |
| Median anti-N IgG, (IQR) | 1.12 (1.29) | 0.87 (1.09) | 0.98 |
| Anti-N IgG, range | 0.07 – 1.71 | 0.06 – 1.82 | --- |
| Median anti-Spike IgG, (IQR) | 1.64 (0.29) | 1.64 (0.29) | 0.89 |
| Anti-Spike IgG, range | 1.07 – 1.82 | 1.28 – 1.96 | --- |
| Median anti-RBD IgG, (IQR) | 1.74 (0.12) | 1.74 (0.12) | 0.82 |
| Anti-RBD IgG, range | 0.70 – 3.03 | 0.74 – 1.93 | --- |
| Median %Neutralization, (IQR) | 98.69 (4.62) | 87.96 (35.28) | 0.06 |
| %Neutralization, range | 7.91 – 99.82 | 0.00 – 99.75 | --- |
| **Neutralizing efficiency, (%)** | --- | --- | --- |
| Neutralization ≤ 30% (weak to negative response) | 1 (4.2) | 3 (10.7) | 0.88 |
| >30% Neutralization <85% | 3 (12.5) | 11 (39.3) | REF |
| ≥85% Neutralization (efficient neutralizers) | 20 (83.3) | 14 (50.0) | 0.03 |
| ^a^ Number missing for variables, Cases: PCR test date 0;  ^b^ Number missing for variables, Controls: PCR test date 7;  ^c^ *P* value <0.05: chi-squared and Fisher’s exact tests used for categorical variables; Wilcoxon Rank Sum test used for continuous variables;  ^d^ Signal-to-cutoff ratio | | | |

| **Table S2a: Serological findings of PCC-cases and infected-controls with baseline assessment 14-365 days post COVID-19 infection^b^** | | | |
| --- | --- | --- | --- |
|  | **Cases (n=93)** | **Controls (n=100)** | ***P* value^a^** |
| **Wave of initial infection, (%)**  Wave 1, March 2020 – August 2020  Wave 2, September 2020 – February 2021  Wave 3, March 2021 – August 2021 | ---  45 (48.4)  33 (35.5)  15 (16.1) | ---  35 (35.0)  43 (43.0)  22 (22.0) | **---**  0.12  0.77  REF |
| **Baseline serology results**  Positive call for anti-Spike IgG (SCO >1, %)^c^ | 87 (93.5) | 87 (87.0) | 0.15 |
| Positive call for anti-RBD IgG (SCO >1, %)^c^ | 88 (94.6) | 81 (81.0) | <0.01 |
| Positive call for anti-N IgG (SCO >1, %)^c^ | 63 (67.7) | 66 (66.0) | 0.80 |
| Median anti-N IgG, (IQR) | 1.00 (1.24) | 0.91 (1.17) | 0.57 |
| Anti-N IgG, range | 0.03 – 2.04 | 0.06 – 2.01 | **---** |
| Median anti-Spike IgG, (IQR) | 1.50 (0.61) | 1.42 (0.73) | 0.21 |
| Anti-Spike IgG, range | 0.01 – 2.57 | 0.01 – 2.22 | **---** |
| Median anti-RBD IgG, (IQR) | 1.26 (1.17) | 1.16 (1.35) | 0.52 |
| Anti-RBD IgG, range | 0.00 – 3.20 | 0.00 – 2.73 | **---** |
| Median %Neutralization, (IQR) | 11.4 (82.1) | 11.4 (62.5) | 0.23 |
| %Neutralization, range | 0.00 – 99.8 | 0.00 – 99.7 | **---** |
| **Neutralizing efficiency, (%)** | **---** | **---** | **---** |
| Neutralization ≤ 30% (weak to negative response) | 58 (62.4) | 65 (65.0) | REF |
| >30% Neutralization <85% | 12 (12.9) | 21 (21.0) | 0.27 |
| ≥85% Neutralization (efficient neutralizers) | 23 (24.7) | 14 (14.0) | 0.11 |
| ^a^ *P* value <0.05: chi-squared and Fisher’s exact tests used for categorical variables; Wilcoxon Rank Sum test used for continuous variables;  ^b^Among participants with known date of COVID-19 infection;  ^c^ Signal-to-cutoff ratio | | | |

| **Table S2b: Serological findings of PCC-cases and infected-controls with baseline assessment 14-180 days post COVID-19 infection^b^** | | | |
| --- | --- | --- | --- |
|  | **Cases (n=47)** | **Controls (n=69)** | ***P* value^a^** |
| **Wave of initial infection, (%)**  Wave 1, March 2020 – August 2020  Wave 2, September 2020 – February 2021  Wave 3, March 2021 – August 2021 | ---  5 (10.6)  27 (57.4)  15 (31.9) | ---  11 (15.9)  37 (53.6)  21 (30.4) | **---**  0.48  0.96  REF |
| **Baseline serology results**  Positive call for anti-Spike IgG (SCO >1, %)^c^ | ---  44 (93.6) | ---  56 (81.2) | ---  0.10 |
| Positive call for anti-RBD IgG (SCO >1, %)^c^ | 45 (95.7) | 54 (78.3) | 0.01 |
| Positive call for anti-N IgG (SCO >1, %)^c^ | 33 (70.2) | 46 (66.7) | 0.69 |
| Median anti-N IgG, (IQR) | 1.23 (1.32) | 1.07 (1.26) | 0.69 |
| Anti-N IgG, range | 0.07 – 1.73 | 0.07 – 2.01 | **---** |
| Median anti-Spike IgG, (IQR) | 1.50 (0.50) | 1.36 (1.04) | 0.06 |
| Anti-Spike IgG, range | 0.00 – 2.22 | 0.01 – 2.22 | **---** |
| Median anti-RBD IgG, (IQR) | 1.47 (1.05) | 1.13 (1.35) | 0.01 |
| Anti-RBD IgG, range | 0.00 – 2.85 | 0.00 – 2.70 | **---** |
| Median %Neutralization, (IQR) | 28.1 (91.7) | 13.4 (65.5) | 0.10 |
| %Neutralization, range | 0.00 – 99.8 | 0.00 – 99.8 | **---** |
| **Neutralizing efficiency, (%)** | **---** | **---** | **---** |
| Neutralization ≤ 30% (weak to negative response) | 25 (53.2) | 42 (60.9) | REF |
| >30% Neutralization <85% | 6 (12.8) | 15 (21.7) | 0.47 |
| ≥85% Neutralization (efficient neutralizers) | 16 (34.0) | 12 (17.4) | 0.08 |
| ^a^ *P* value <0.05: chi-squared and Fisher’s exact tests used for categorical variables; Wilcoxon Rank Sum test used for continuous variables; ^b^Among participants with known date of COVID-19 infection; ^c^ Signal-to-cutoff ratio **Table S2c: Serological findings of PCC-cases and infected-controls with baseline assessment 14-90 days post COVID-19 infection^b^** \| \| \|; \| **Cases (n=23)** \| **Controls (n=45)** \| ***P* value^a^**; **Wave of initial infection, (%)** Wave 1, March 2020 – August 2020 Wave 2, September 2020 – February 2021 Wave 3, March 2021 – August 2021 \| **---** 1 (4.3) 16 (69.6) 6 (26.1) \| **---** 2 (4.4) 33 (73.3) 10 (22.2) \| **---** 0.89 0.72 REF; **Baseline serology results** Positive call for anti-Spike IgG (SCO >1, %)^c^ \| --- 21 (91.3) \| --- 35 (77.8) \| --- 0.20; Positive call for anti-RBD IgG (SCO >1, %)^c^ \| 22 (95.7) \| 34 (75.6) \| 0.05; Positive call for anti-N IgG (SCO >1, %)^c^ \| 16 (69.6) \| 30 (66.7) \| 1.00; Median anti-N IgG, (IQR) \| 1.39 (1.36) \| 1.07 (1.26) \| 0.82; Anti-N IgG, range \| 0.07 – 1.73 \| 0.07 – 2.01 \| **---**; Median anti-Spike IgG, (IQR) \| 1.44 (0.68) \| 1.41 (1.06) \| 0.17; Anti-Spike IgG, range \| 0.00 – 1.89 \| 0.01 – 2.22 \| **---**; Median anti-RBD IgG, (IQR) \| 0.92 (1.17) \| 1.03 (1.40) \| 0.04; Anti-RBD IgG, range \| 0.00 – 2.05 \| 0.00 – 2.70 \| **---**; Median %Neutralization, (IQR) \| 11.27 (28.37) \| 6.33 (44.40) \| 0.61; %Neutralization, range \| 0.00 – 97.0 \| 0.00 – 99.4 \| **---**; **Neutralizing efficiency, (%)** \| **---** \| **---** \| **---**; Neutralization ≤ 30% (weak to negative response) \| 18 (78.3) \| 31 (68.9) \| REF; >30% Neutralization <85% \| 3 (13.0) \| 11 (24.4) \| 0.29; ≥85% Neutralization (efficient neutralizers) \| 2 (8.7) \| 3 (6.7) \| 0.89; ^a^ *P* value <0.05: chi-squared and Fisher’s exact tests used for categorical variables; Wilcoxon Rank Sum test used for continuous variables; ^b^Among participants with known date of COVID-19 infection; ^c^ Signal-to-cutoff ratio \| \| \| | | | |

| **FIGURE S1: Receiving operating characteristic (ROC) curves for minimally adjusted (LEFT) and fully adjusted (RIGHT) models** \| \|; **Effect of anti-Spike IgG titres on odds of PCC,** **adjusted for age and sex** \| \| **Effect of anti-Spike IgG titres on odds of PCC, adjusted for age, sex, allergies, need for hospitalization/medical help for COVID-19 symptoms, and time since infection**; 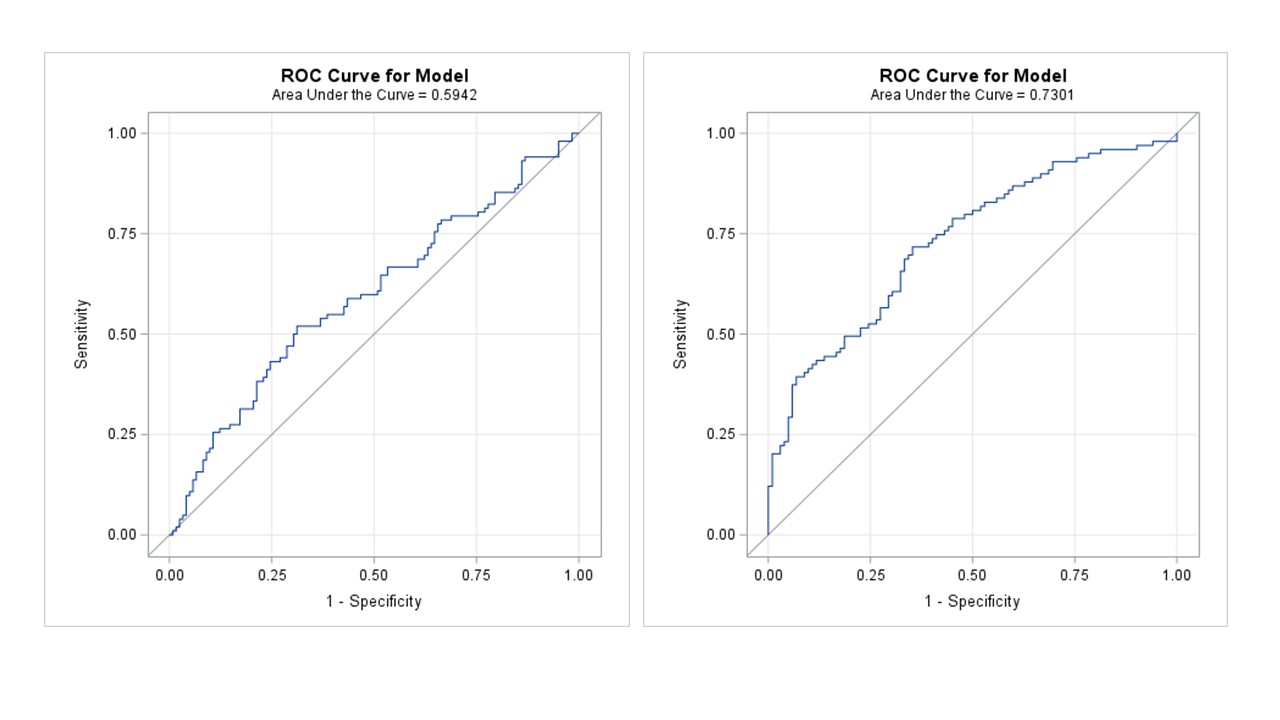 \| \|; **Effect of anti-RBD IgG titres on odds of PCC,** **adjusted for age and sex** \| \| **Effect of anti-RBD IgG titres on odds of PCC, adjusted for age, sex, allergies, need for hospitalization/medical help for COVID-19 symptoms, and time since infection**; 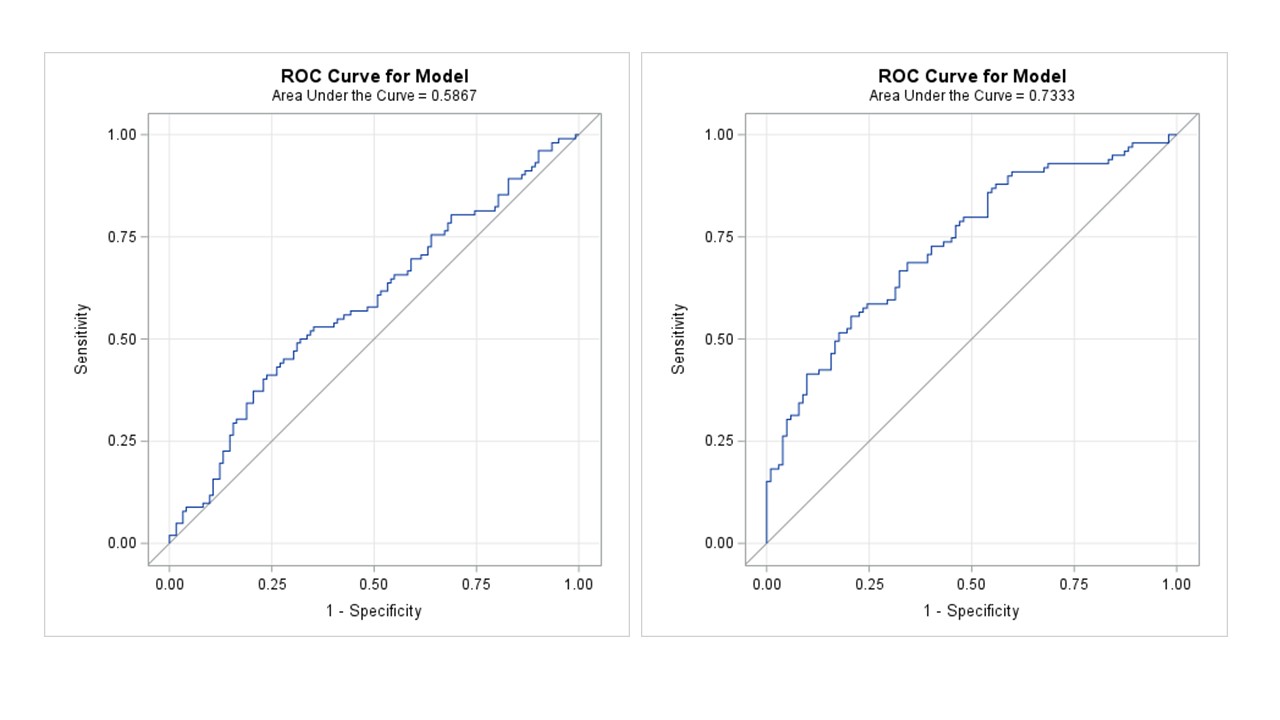 \| \|; **Effect of anti-N IgG titres on odds of PCC,** **adjusted for age and sex** \| \| **Effect of anti-N IgG titres on odds of PCC, adjusted for age, sex, allergies, need for hospitalization/medical help for COVID-19 symptoms, and time since infection**; 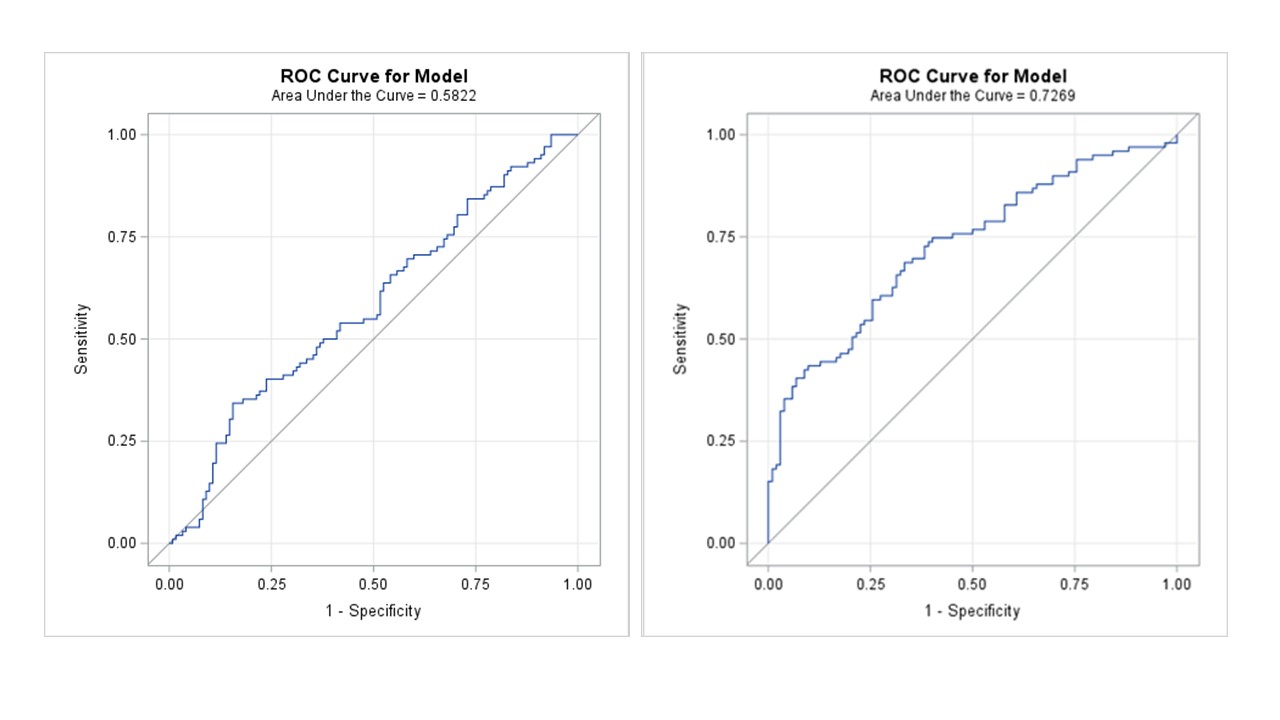 \| \|; **Effect of efficient neutralization on odds of PCC,** **adjusted for age and sex** \| \| **Effect of efficient neutralization on odds of PCC, adjusted for age, sex, allergies, need for hospitalization/medical help for COVID-19 symptoms, and time since infection**; 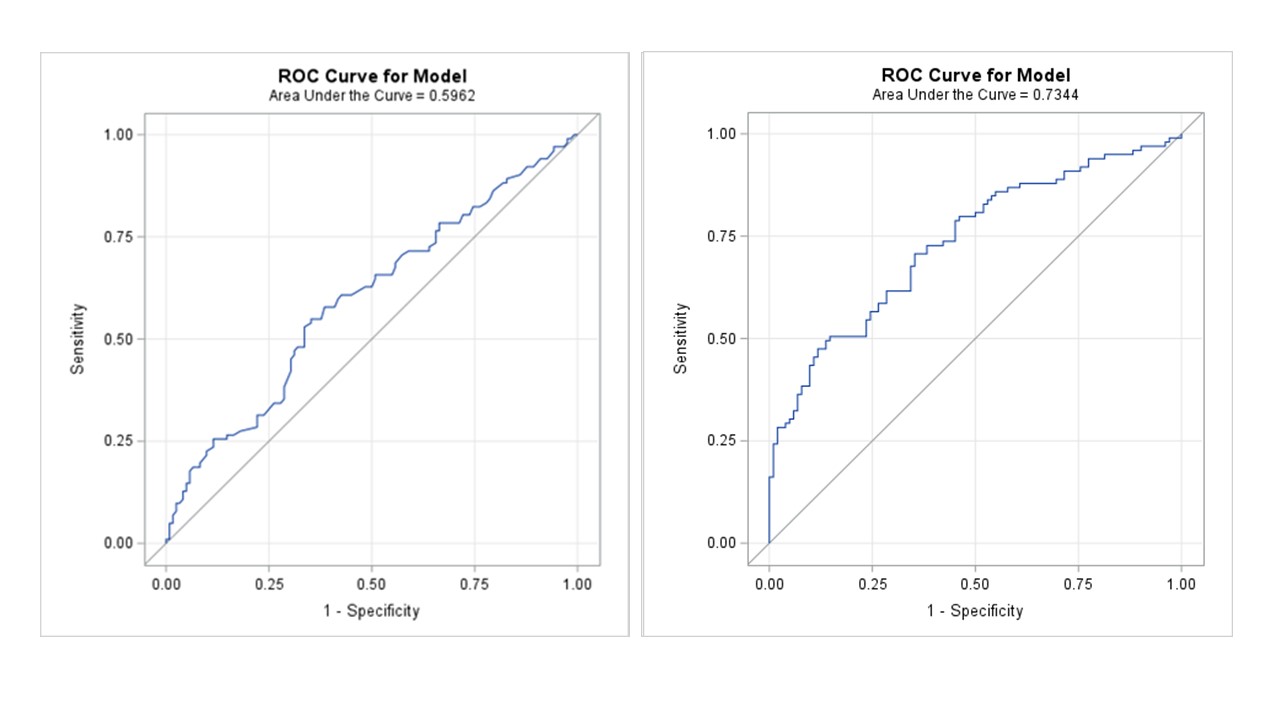 \| \| | | | |
| --- | --- | --- | --- |
| **TABLE S3a: Effect of anti-Spike IgG titres transformed with restricted cubic splines (k=3, percentiles=10, 50, 90) on odds of Post COVID-19 Condition (PCC) in minimally and fully adjusted models** | | | |
|  | **Unadjusted** | **Minimally adjusted model^d^** | **Fully adjusted model^e^** |
| **OR (95% CI)** | **---** | **---** | **---** |
| Anti-Spike IgG^a^ | 1.2 (0.81 – 1.91) | 1.2 (0.79 – 1.91) | 1.2 (0.67 – 1.97) |
| Sex, male | 0.7 (0.43 – 1.26) | 0.7 (0.38 – 1.17) | 1.0 (0.51 – 1.81) |
| Age^b^ | 1.3 (0.86 – 2.10) | 1.3 (0.83 – 2.12) | 1.2 (0.68 – 1.99) |
| Time since infection, months^c^ | 2.6 (1.46 – 4.45) | **---** | 2.0 (1.07 – 3.55) |
| Sought medical help or hospitalized for  COVID-19 symptoms | ---  4.1 (2.26 – 7.38) | **---**  **---** | ---  3.3 (1.67 – 6.44) |
| Allergies | 1.8 (1.04 – 3.00) | **---** | 1.8 (0.98 – 3.44) |
| ***P* value for trend** | --- | 0.41 | 0.78 |
| **C-statistic** | --- | 0.59 | 0.73 |
| **Adjusted R^2^** | --- | 0.03 | 0.21 |
| **Bayesian information criterion (BIC)** | --- | 335.77 | 297.04 |
| ^a^RCS-transformed, effect on odds of PCC given change from first to third quartile (1.03 – 1.71 anti-Spike SLUs)  ^b^RCS-transformed, effect on odds of PCC given change from first to third quartile (34.8 – 59.0 years)  ^c^RCS-transformed, effect on odds of PCC given change from first to third quartile (5.0 – 18.7 months)  ^d^Adjusted for age and sex  ^e^Adjusted for age, sex, allergies, hospitalized or sought medical help for COVID-19 symptoms, and time since infection | | | |
| **TABLE S3b: Effect of anti-N IgG titres transformed with restricted cubic splines (k=3, percentiles=10, 50, 90) on odds of Post COVID-19 Condition (PCC) in minimally and fully adjusted models** | | | |
|  | **Unadjusted** | **Minimally adjusted model^d^** | **Fully adjusted model^e^** |
| **OR (95% CI)** | **---** | **---** | **---** |
| Anti-N IgG^a^ | 1.1 (0.63 – 1.82) | 1.0 (0.58 – 1.80) | 1.2 (0.64 – 2.39) |
| Sex, male | 0.7 (0.43 – 1.26) | 0.7 (0.39 –1.20) | 0.9 (0.49 – 1.76) |
| Age^b^ | 1.3 (0.86 – 2.10) | 1.4 (0.87 – 2.25) | 1.3 (0.7 – 1.98) |
| Time since infection, months^c^ | 2.6 (1.46 – 4.45) | **---** | 2.1 (1.14 – 3.79) |
| Sought medical help or hospitalized for  COVID-19 symptoms | ---  4.1 (2.26 – 7.38) | **---**  **---** | ---  3.3 (1.67 – 6.44) |
| Allergies | 1.8 (1.04 – 3.00) | **---** | 1.8 (0.97 – 3.34) |
| ***P* value for trend** | --- | 0.61 | 0.80 |
| **C-statistic** | --- | 0.58 | 0.73 |
| **Adjusted R^2^** | --- | 0.03 | 0.21 |
| **Bayesian information criterion (BIC)** | --- | 336.61 | 297.08 |
| ^a^RCS-transformed, effect on odds of PCC given change from first to third quartile (0.33 – 1.53 anti-N SLUs)  ^b^RCS-transformed, effect on odds of PCC given change from first to third quartile (34.8 – 59.0 years)  ^c^RCS-transformed, effect on odds of PCC given change from first to third quartile (5.0 – 18.7 months)  ^d^Adjusted for age and sex  ^e^Adjusted for age, sex, allergies, hospitalized or sought medical help for COVID-19 symptoms, and time since infection | | | |
| **TABLE S3c: Effect of anti-RBD IgG titres transformed with restricted cubic splines (k=3, percentiles=10, 50, 90) on odds of Post COVID-19 Condition (PCC) in minimally and fully adjusted models** | | | |
|  | **Unadjusted** | **Minimally adjusted model^d^** | **Fully adjusted model^e^** |
| **OR (95% CI)** | **---** | **---** | **---** |
| Anti-RBD IgG^a^ | 1.2 (0.75 – 1.92) | 1.1 (0.70 - 1.83) | 1.1 (0.64 - 1.88) |
| Sex, male | 0.7 (0.43 – 1.26) | 0.7 (0.39 – 1.20) | 1.0 (0.52 - 1.86) |
| Age^b^ | 1.3 (0.86 – 2.10) | 1.5 (0.92 - 2.40) | 1.3 (0.76 – 2.28) |
| Time since infection, months^c^ | 2.6 (1.46 – 4.45) | **---** | 2.0 (1.11 – 3.63) |
| Sought medical help or hospitalized for  COVID-19 symptoms | ---  4.1 (2.26 – 7.38) | **---**  **---** | ---  3.4 (1.71 - 6.60) |
| Allergies | 1.8 (1.04 – 3.00) | **---** | 1.8 (0.97 – 3.31) |
| ***P* value for trend** | --- | 0.45 | 0.40 |
| **C-statistic** | --- | 0.59 | 0.73 |
| **Adjusted R^2^** | --- | 0.03 | 0.22 |
| **Bayesian information criterion (BIC)** | --- | 336.20 | 295.82 |
| ^a^RCS-transformed, effect on odds of PCC given change from first to third quartile (0.45 – 1.75 anti-RBD SLUs)  ^b^RCS-transformed, effect on odds of PCC given change from first to third quartile (34.8 – 59.0 years)  ^c^RCS-transformed, effect on odds of PCC given change from first to third quartile (5.0 – 18.7 months)  ^d^Adjusted for age and sex  ^e^Adjusted for age, sex, allergies, hospitalized or sought medical help for COVID-19 symptoms, and time since infection | | | |
| **TABLE S3d: Effect of efficient neutralization (%Neutralization ≥85%) on odds of Post COVID-19 Condition (PCC) in minimally and fully adjusted models** | | | |
|  | **Unadjusted** | **Minimally adjusted model^c^** | **Fully adjusted model^d^** |
| **OR (95% CI)** | **---** | **---** | **---** |
| %Neutralization ≥85% | 2.3 (1.14 – 4.51) | 2.2 (1.11 – 4.49) | 2.0 (0.89 – 4.54) |
| Sex, male | 0.7 (0.43 – 1.26) | 0.7 (0.42 – 1.30) | 1.0 (0.54 – 1.96) |
| Age^a^ | 1.3 (0.86 – 2.10) | 1.4 (0.90 – 2.28) | 1.2 (0.72 – 2.08) |
| Time since infection, months^b^ | 2.6 (1.46 – 4.45) | **---** | 1.9 (1.06 – 3.51) |
| Sought medical help or hospitalized for  COVID-19 symptoms | ---  4.1 (2.26 – 7.38) | **---**  **---** | ---  3.2 (1.61 – 6.24) |
| Allergies | 1.8 (1.04 – 3.00) | **---** | 1.9 (1.00 – 3.45) |
| **C-statistic** | --- | 0.60 | 0.73 |
| **Adjusted R^2^** | --- | 0.05 | 0.22 |
| **Bayesian information criterion (BIC)** | --- | 326.94 | 289.39 |
| ^a^RCS-transformed, effect on odds of PCC given change from first to third quartile (34.8 – 59.0 years)  ^b^RCS-transformed, effect on odds of PCC given change from first to third quartile (5.0 – 18.7 months)  ^c^Adjusted for age and sex  ^d^Adjusted for age, sex, allergies, hospitalized or sought medical help for COVID-19 symptoms, and time since infection | | | |
